# SARS-CoV2 mRNA-vaccination-induced Immunological Memory in Human Non-Lymphoid and Lymphoid Tissues

**DOI:** 10.1101/2023.02.22.23286293

**Authors:** Vanessa Proß, Arne Sattler, Sören Lukassen, Laura Tóth, Linda Marie Laura Thole, Janine Siegle, Carolin Stahl, An He, Georg Damm, Daniel Seehofer, Christina Götz, Christian Bayerl, Pia Jäger, Alexander Macke, Stephan Eggeling, Bernadette Kirzinger, Thomas Mayr, Hermann Herbst, Katharina Beyer, Dominik Laue, Jan Krönke, Jan Braune, Friederike Rosseck, Frank Friedersdorff, Mandy Hubatsch, Sarah Weinberger, Nils Lachmann, Veit Maria Hofmann, Eva Schrezenmeier, Carolin Ludwig, Hubert Schrezenmeier, Katharina Jechow, Christian Conrad, Katja Kotsch

**Affiliations:** Department of General- and Visceral Surgery, Charité – Universitätsmedizin Berlin, corporate member of Freie Universität Berlin, Humboldt-Universität zu Berlin, and Berlin Institute of Health, Berlin, Germany; Center for Digital Health, Berlin Institute of Health (BIH) and Charité - Universitätsmedizin Berlin, corporate member of Freie Universität Berlin, Humboldt-Universität zu Berlin, Berlin, Germany; Department of Hepatobiliary Surgery and Visceral Transplantation, University Hospital, Leipzig University, Leipzig, Germany; Department of Radiology, Charité - Universitätsmedizin Berlin, Campus Benjamin Franklin, Corporate Member of Freie Universität Berlin and Humboldt-Universität zu Berlin, and Berlin Institute of Health, Berlin, Germany; Department of Visceral Surgery, Vivantes Klinikum Neukölln, Berlin, Germany; Department of Thoracic Surgery, Vivantes Klinikum Neukölln, Berlin, Germany; Department of Pathology, Vivantes Klinikum Neukölln, Berlin, Germany; Department of Traumatology and Reconstructive Surgery, Campus Benjamin Franklin, Charité-Universitätsmedizin Berlin, corporate member of Freie Universität Berlin, Humboldt-Universität zu Berlin, and Berlin Institute of Health, Berlin, Germany; Department of Hematology, Oncology and Cancer Immunology, Charité - Universitätsmedizin Berlin, corporate member of Freie Universität Berlin and Humboldt-Universität zu Berlin, Berlin, Germany; Institute of Pathology, Charité-Universitätsmedizin Berlin, Corporate Member of Freie Universität Berlin, Humboldt-Universität zu Berlin and Berlin Institute of Health, Berlin, Germany; Department of Urology, Evangelisches Krankenhaus Königin Elisabeth Herzberge, Berlin, Germany; Department of Urology, Charité-Universitätsmedizin Berlin, corporate member of Freie Universität Berlin, Humboldt-Universität zu Berlin, and Berlin Institute of Health; Institute of Transfusion Medicine, Berlin Institute of Health, Charité - Universitätsmedizin Berlin, Humboldt-Universität zu Berlin, Berlin, Germany; Department of Otolaryngology, Charité-Universitätsmedizin Berlin, corporate member of Freie Universität Berlin, Humboldt-Universität zu Berlin, and Berlin Institute of Health; Department of Nephrology and Medical Intensive Care, Charité-Universitätsmedizin Berlin, Corporate Member of Freie Universität Berlin and Humboldt-Universität zu Berlin, Berlin, Germany; BIH Charité Clinician Scientist Program, BIH Biomedical Innovation Academy, Berlin Institute of Health at Charité–Universita□tsmedizin Berlin, Berlin, Germany; Institute for Clinical Transfusion Medicine and Immunogenetics, German Red Cross Blood Transfusion Service Baden-Württemberg-Hessen and University Hospital Ulm, Ulm, Germany

## Abstract

Tissue-resident lymphocytes provide organ-adapted protection against invading pathogens. Whereas their biology has been examined in great detail in various infection models, their generation and functionality in response to vaccination has not been comprehensively analyzed in humans. We therefore studied SARS-CoV2 mRNA-vaccine-specific T cells in surgery specimens of kidney, liver, lung, bone marrow and spleen in comparison to paired blood samples from largely virus-naïve individuals. As opposed to lymphoid tissues, non-lymphoid organs harbored significantly elevated frequencies of Spike-specific CD4^+^ T cells compared to paired peripheral blood showing hallmarks of tissue residency and an expanded memory pool. Organ-derived, vaccine-specific T helper (Th) cells were characterized by increased portions of multifunctional cells over those detected in blood. Single-cell RNA sequencing revealed functional rather than organ-specific clusters of Spike-reactive Th cells, indicating similar diversification programs across tissues. T cell receptor (TCR) repertoire analysis indicated that the TCR sequence is a major determinant of transcriptomic state in tissue-resident, vaccine-specific CD4^+^ T cells. In summary, our data demonstrate that SARS-CoV2 vaccination entails acquisition of tissue memory and residency features in organs distant from the inoculation site, thereby contributing to our understanding of how local tissue protection might be accomplished.

**One sentence summary:** SARS-CoV2 mRNA vaccination-induced CD4^+^ Th cells reside in both human lymphoid and non-lymphoid organs showing distinct adaptations in tissues with respect to memory differentiation, retention and function.

## Introduction

Vaccination against SARS-CoV2 has been proven in large cohort studies (*1, 2*) a powerful strategy to protect from severe corona virus disease 2019 (COVID-19). Since the availability of the first SARS-CoV2 vaccines in late 2020, specific humoral and cellular immunity has been characterized in unpreceded depth in healthy individuals and those with multiple pre-conditions or comorbidities (*3-5*). Whereas infection- or vaccination-induced T cell biology has been comprehensively examined in peripheral blood, only limited information is available as to how immunological memory is established in tissues. Postulated more than 10 years ago, the concept of tissue residency has been raised in context with different tissue-targeted infections, implying that a substantial portion of memory T cells has a limited capacity to re-circulate, but acquires residency in organs, thus providing site-adapted immunity (*6-8*). Pioneering work noted a distribution of SARS-CoV2 infection-induced T cells across multiple human tissues, including lymph nodes, spleen, lung and bone marrow. Importantly, a tissue residency signature, as reflected by CD69 and CD103 expression, was mainly identified in lung, but absent in bone marrow (BM) (*9*). Multi-organ residency is also characteristic for cytomegalovirus (CMV) specific T cells (*10*) that have been recently detected in high frequencies, along with those specific for Epstein-Barr virus (EBV-) and influenza, in resected human kidneys (*11*).

With respect to features of vaccination-induced T cell memory, most comprehensive analyses in organs have been conducted using human bone marrow where CD4^+^ T cells specific for multiple vaccination-associated antigens, including tetanus toxoid, measles and mumps have been characterized (*12*). In comparison to peripheral blood, CD4^+^ T helper (Th) cells were in a resting state and upregulated CD69, both being indicative for bone marrow as a niche for long-term maintenance of resident memory for systemic pathogens (*13*). So far, it remains to be determined how SARS-CoV2 mRNA vaccination-induced T cell memory is maintained with respect to organ tropism and tissue adaptation, particularly considering non-lymphoid organs. Both features might be potentially influenced by *in vivo* distribution of vaccination antigens. For novel mRNA-based vaccines, distribution and degradation of encoded proteins have been followed in experimental models. After intramuscular application, protein expression was predominantly confined to the injection site, but also included distal organs such as lung and liver, indicating systemic spread of mRNA containing lipid nanoparticles (*14*). Similar findings on biodistribution were made for a vector-based SARS-CoV2 vaccine (AZD1222/ChAdOx1) that was detectable for up to 29 days in bone marrow, liver, lung, spleen and lymph nodes (*15*). It is therefore principally conceivable that differentiation and/or recruitment of cellular immunity involves organs distant to the vaccination site. To address tissue distribution of SARS-CoV2 vaccine-specific T cell memory, we examined human lymphoid (bone marrow, spleen, tonsil) and non-lymphoid (liver, kidney, lung) organs for quantities and functional features of Spike protein-specific T cells in comparison with paired blood samples. Here, we identify mRNA vaccine-specific CD4^+^ T cells in most tissue types examined with distinct adaptations particularly identified for non-lymphoid organs.

## Results

### Donor tissue cohort

In order to analyze SARS-CoV2 vaccine-specific T- and B cells in various human tissues, specimens of solid, non-lymphoid (liver, kidney, lung) as well as lymphoid (bone marrow, spleen, tonsil) organs were procured together with paired blood samples (Figure 1A) between October 2021 and October 2022. Surgeries were primarily, but not exclusively conducted for tumor resection; in these cases, peri-tumor tissue located most distant to the tumor was used unless otherwise indicated. To focus on vaccination-induced immunity, the majority of individuals enrolled was SARS-CoV2 naïve as evidenced by medical history and absence of reactivity in a SARS-CoV2 nucleocapsid-specific ELISA. Details on patient demographics, including type and time since last vaccination are summarized in Table 1. Tissue-, blood- and serum samples were immediately processed after collection and cryopreserved before assessment of vaccine-specific immunity as summarized in Figure 1B.Depending on the time point of sample procurement, vaccination history of individuals comprised two or three mRNA vaccine doses (BioNTech/Pfizer or Moderna) (Table 1).

**Table 1.** Demographics of patients enrolled.

| Variable |  | Age<br>min/max<br>(yrs) | Mean age<br>(years±SD) | Vaccinated +<br>infected (%) |
| --- | --- | --- | --- | --- |
| **Total number of patients (%)** | 61 (100) |  |  |  |
| **Sex** (male/female/ns) | 32 (52.4)/25 (40.9)<br>4 (6.5) |  |  |  |
| **Cause of surgery:** |  |  |  |  |
| **Liver (n=12):** |  | 28/79 | 60.3±15.1 | 1 (8.3) |
| Hepatocellular carcinoma (%) | 1 (8.3) |  |  |  |
| Colorectal liver metastases (%) | 3 (25.0) |  |  |  |
| Intrahepatic cholangiocarcinoma (%) | 5 (41.6) |  |  |  |
| Liver adenoma (%) | 3 (25.0) |  |  |  |
| Vaccine: Moderna/Biontech (%) | 2 (16.6)/10 (83.3) |  |  |  |
| Time since last vaccination<br>(mean in days±SD) | 118.8±34.1 |  |  |  |
| **Kidney (n=14):** |  | 51/87 | 71.4±10.6 | 0 (0) |
| Renal cell carcinoma (RCC) (%) | 11 (78.5) |  |  |  |
| Urethral cancer (%) | 2 (14.2) |  |  |  |
| Pyelonephroma (%) | 1 (7.14) |  |  |  |
| Vaccine: Moderna/Biontech (%) | 7 (50.0)/7 (50.0) |  |  |  |
| Time since last vaccination<br>(mean in days±SD) | 133.1±75.4 |  |  |  |
| **Lung (n=15):** |  | 58/85 | 69.5±7.8 | 2 (13.3) |
| Non-small/small cell lung cancer (%) | 15 (100.0) |  |  |  |
| Vaccine: Moderna/Biontech (%) | 3 (20.0)/12 (80.0) |  |  |  |
| Time since last vaccination<br>(mean in days±SD) | 92.1±60.1 |  |  |  |
| **Bone marrow (n=11):** |  | 36/78 | 57.8±13.8 | 0(0) |
| Orthopedic spine diseases (%) | 11 (100.0) |  |  |  |
| Vaccine: Moderna/Biontech (%) | 1 (9.1)/10 (90.9) |  |  |  |
| Time since last vaccination<br>(mean in days±SD) | 78.5±68.2 |  |  |  |
| **Spleen (n=6):** |  | 47/74 | 59.0±8.8 | 2(33.3) |
| Splenectomy<br>(oncologic surgery) (%) | 6 (100.0) |  |  |  |
| Vaccine: Moderna/Biontech (%) | 0 (0)/6 (100) |  |  |  |
| Time since last vaccination<br>(mean in days±SD) | 101.2±21.0 |  |  |  |
| **Tonsil (n=3):** |  | 21/31 | 26.3±5.0 | 1(33.3) |
| Acute or chronic tonsillitis (%) | 3 (100.0) |  |  |  |
| Vaccine: Moderna/Biontech (%) | 1 (33.3)/2 (66.6) |  |  |  |
| Time since last vaccination<br>(mean in days±SD) | 129.7±32.0 |  |  |  |
| **Metastases diagnosed before surgery:** |  |  |  |  |
| Distant metastases (%) | 6 (9.8) |  |  |  |
| **Chemotherapy <8 weeks before/after**<br>**vaccination or analysis:** |  |  |  |  |
| Within all patients | 3 (4.9) |  |  |  |
| Within CD4<sup>+</sup> T cell responders | 1 (1.6) |  |  |  |

**Figure 1.**
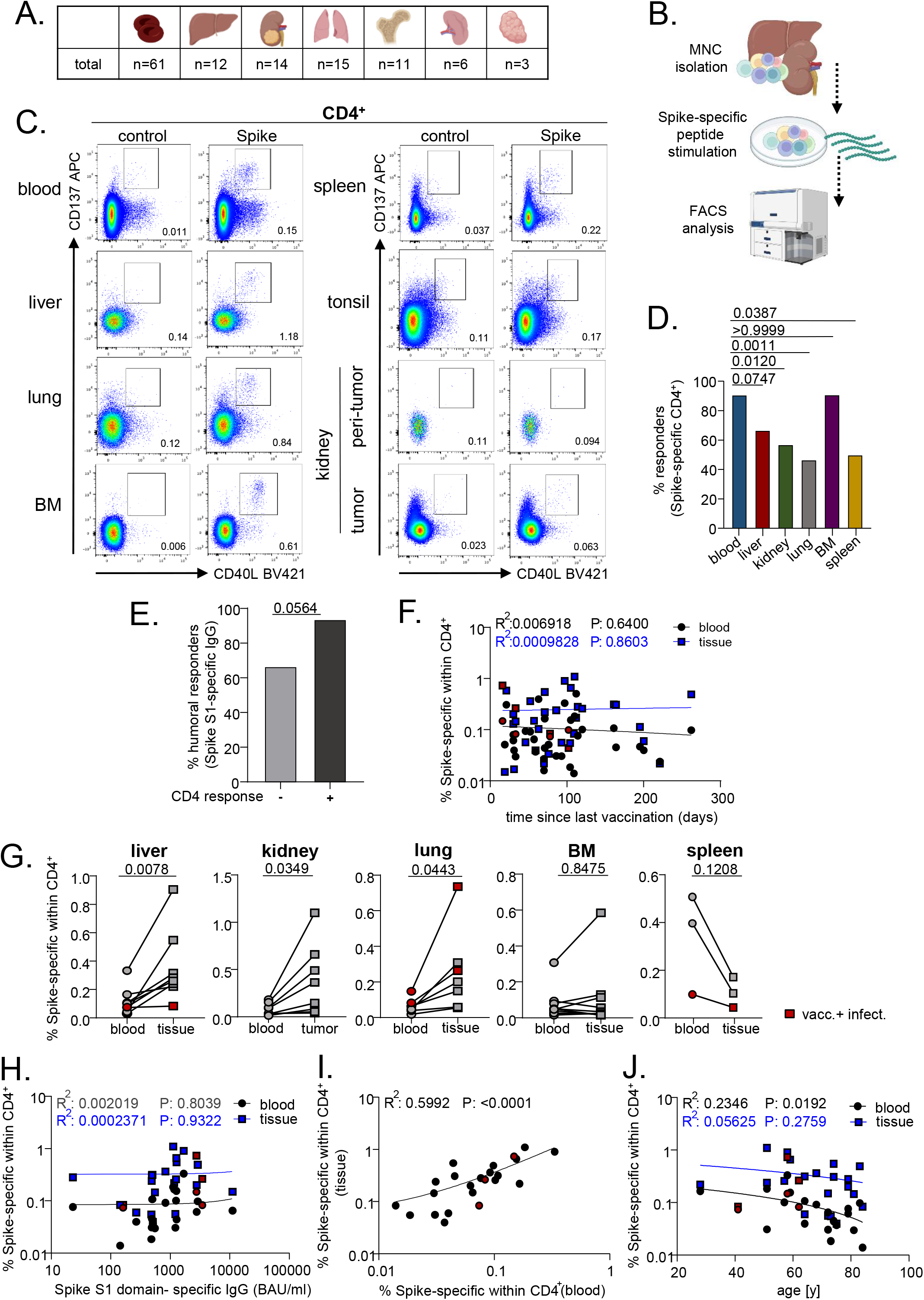
Quantification of SARS-CoV2 vaccine-induced CD4^+^ Th cells in non-lymphoid and lymphoid organs. (A) Summary of all specimens included for analysis of vaccine-specific T cells. (B) Schematic workflow overview. (C) Exemplary plots showing vaccine-specific CD137^+^CD40L^+^ CD4^+^ T cells from the indicated organs as identified by FACS. (D) Portions of individuals showing Spike-specific CD4^+^ T cell responses within the depicted organs. Statistically significant differences were tested with two-sided Fisher’s exact test with n as in (A). (E) Portions of individuals with Spike S1-domain-specific IgG responses, stratified for cellular responders and non-responders. Statistically significant differences were tested with two-sided Fisher’s exact test. (F) Simple linear regression analysis between frequencies of Spike-specific Th cells and time since last vaccination with n as in (A). (G) Pairwise comparison of Spike-specific CD4^+^ T cell frequencies in peripheral blood and organ-derived specimens as indicated. Liver: n=8, Wilcoxon test; kidney: n=8, paired t test; lung: n=7, paired t test; BM: n=10, Wilcoxon test; spleen: n=3, paired t test. (H) Simple linear regression analysis between frequencies of specific Th cells in non-lymphoid organs and Spike S1 domain-specific IgG levels or (I) paired blood samples. (J) Simple linear regression analysis between specific blood- and paired non-lymphoid organ-derived T cell frequencies with age. Red symbols identify vaccinated individuals with a history of SARS-CoV2 infection.

### SARS-CoV-2 vaccine–specific T cells in non-lymphoid and lymphoid tissues

Specific CD4^+^ T helper cells were identified based on CD137 and CD40L co-expression after stimulation with an overlapping peptide pool encompassing the complete SARS-CoV2 Spike protein as outlined in Supplemental Figure 1A. To assess *ex vivo* expressing-, but not stimulation-induced CD69^+^ T cells, mononuclear cells (MNC) were stained with CD69 BV785 before culture; labelling was stable without appreciable loss of signal intensity until stimulation termination as recently demonstrated (*11*) and depicted in Supplemental Figure 1B. Overall, Spike-specific CD4^+^ T cells could be identified in peripheral blood, liver, lung, BM, spleen and kidney tumor-but not in kidney peri-tumor tissue or tonsil as exemplified in Figure 1C. Amongst all samples, cellular responses were most frequently detected in blood and BM (Figure 1D). Individuals with cellular responses in peripheral blood showed a trend towards an elevated rate of specific IgG responses over cellular non-responders (Figure 1E). Overall frequencies of Spike-specific Th cells (Figure 1F) and those exhibiting a memory or effector phenotype or expressing IFNγ or IL-2 (Supplemental Figure 2A) remained constant in blood and tissue with progressing time from last vaccination. Similarly, no significant differences were identified with respect to cellular responder rates (Supplemental Figure 2B), frequencies, memory differentiation or function (Supplemental Figure C) for tumor- vs. non-tumor patient derived blood samples, thereby excluding an appreciable impact of patient preconditions.

For subsequent analyses, comparisons between paired specimens were conducted when both blood and tissue samples fulfilled the criteria for a cellular response as defined in the Methods section. As a common motif, non-lymphoid (liver, kidney tumor, lung), but not lymphoid (BM, spleen) tissues were characterized by significantly elevated frequencies of Spike-specific CD4^+^ T cells over those quantified in blood (Figure 1G). We did not note a significant correlation between Spike-specific IgG levels and frequencies of blood- or non-lymphoid tissue-derived CD4^+^ T cells (Figure 1H). However, frequencies of specific CD4^+^ T 7 cells detected in blood significantly correlated with those in paired non-lymphoid organs (Figure 1I) and showed a trend for lymphoid organs (Supplemental Figure 3A). Interestingly, portions of Spike-specific CD4^+^ T cells significantly declined with age in blood, but not in non-lymphoid tissues (Figure 1J); this observation did not apply to lymphoid tissues (Supplemental Figure 3B). In summary, vaccine-specific CD4^+^ Th cell responses in largely virus-naïve individuals could be detected both in lymphoid and non-lymphoid organs. Only for the latter, we observed an enrichment of specific cells in tissue-compared to blood-derived specimens.

### Memory differentiation and tissue adaptation of vaccine-specific CD4^+^ T cells

To identify distinct organ-specific adaptation patterns, vaccine-specific CD4^+^ lymphocytes were further characterized according to expression of typical molecules reflecting memory phenotype (CD45RO, CD62L) and tissue residency (CD69, CD103, CD49a). Non-lymphoid organs were enriched for specific CD45RO^+^CD62L^-^ memory-type T cells (T_M_), along with a drop in CD45RO^-^CD62L^-^ effector-type cells (T_EFF_) as compared to paired blood; the T_M_ pattern also showed a similar trend for bone marrow, but not for spleen, also owing to sample size (Figure 2A and B). In contrast to non-lymphoid tissues, frequencies of antigen-specific T_M_ declined in peripheral blood according to age (Figure 2C). This dichotomy was not evident for lymphoid tissue and paired blood specimens (Supplemental Figure 3C). Of note, a similar segregation between both organ systems could be observed based on frequencies of CD69^+^ and CD49a^+^ cells that tended to be or were significantly elevated in non-lymphoid, but not lymphoid organs compared to paired blood samples with the exception of CD69 in spleen (Figure 3A and B). Interestingly, the integrin CD103 was only detectable in a minor portion of antigen-experienced CD4^+^ T cells and expression was mainly confined to the lung.

**Figure 2.**
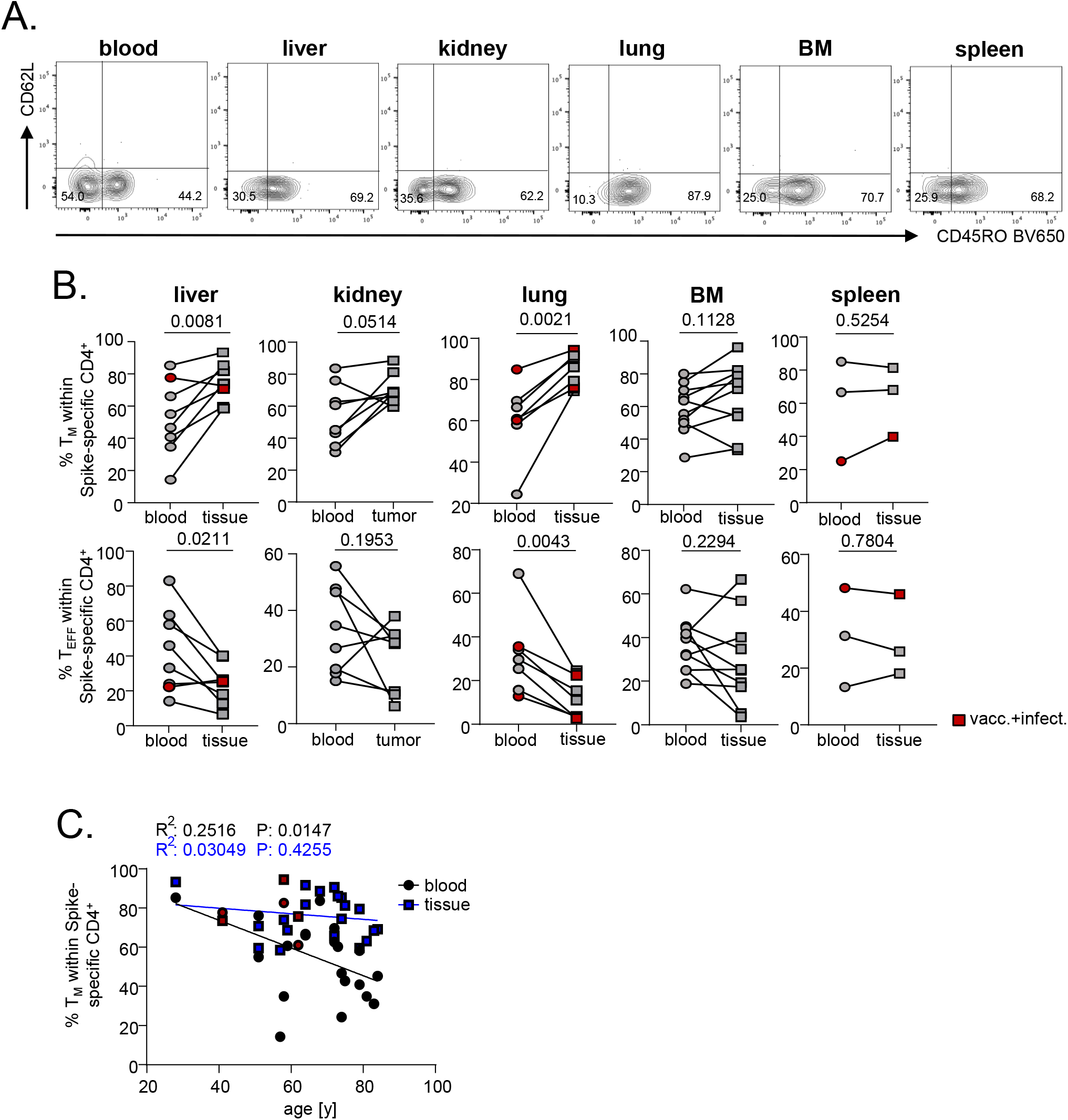
Enrichment of specific memory-type CD4^+^ T cells in non-lymphoid tissues. (A) Exemplary plots for FACS-based identification of CD45RO^+^CD62L^-^ memory- (T_M_) and CD45RO^-^CD62L^-^ effector-type (T_EFF_) T cells within the Spike-specific compartment of different paired samples as summarized in (B). Liver: n=8, paired t test; kidney: n=8, paired t test for T_M_ and Wilcoxon test for T_EFF_; lung: n=7, paired t test; BM: n=10, paired t test; spleen: n=3, paired t test. (C) Simple linear regression analysis between specific blood- and paired non-lymphoid organ-derived T_M_ cell frequencies with age. Red symbols identify vaccinated individuals with a history of SARS-CoV2 infection.

**Figure 3.**
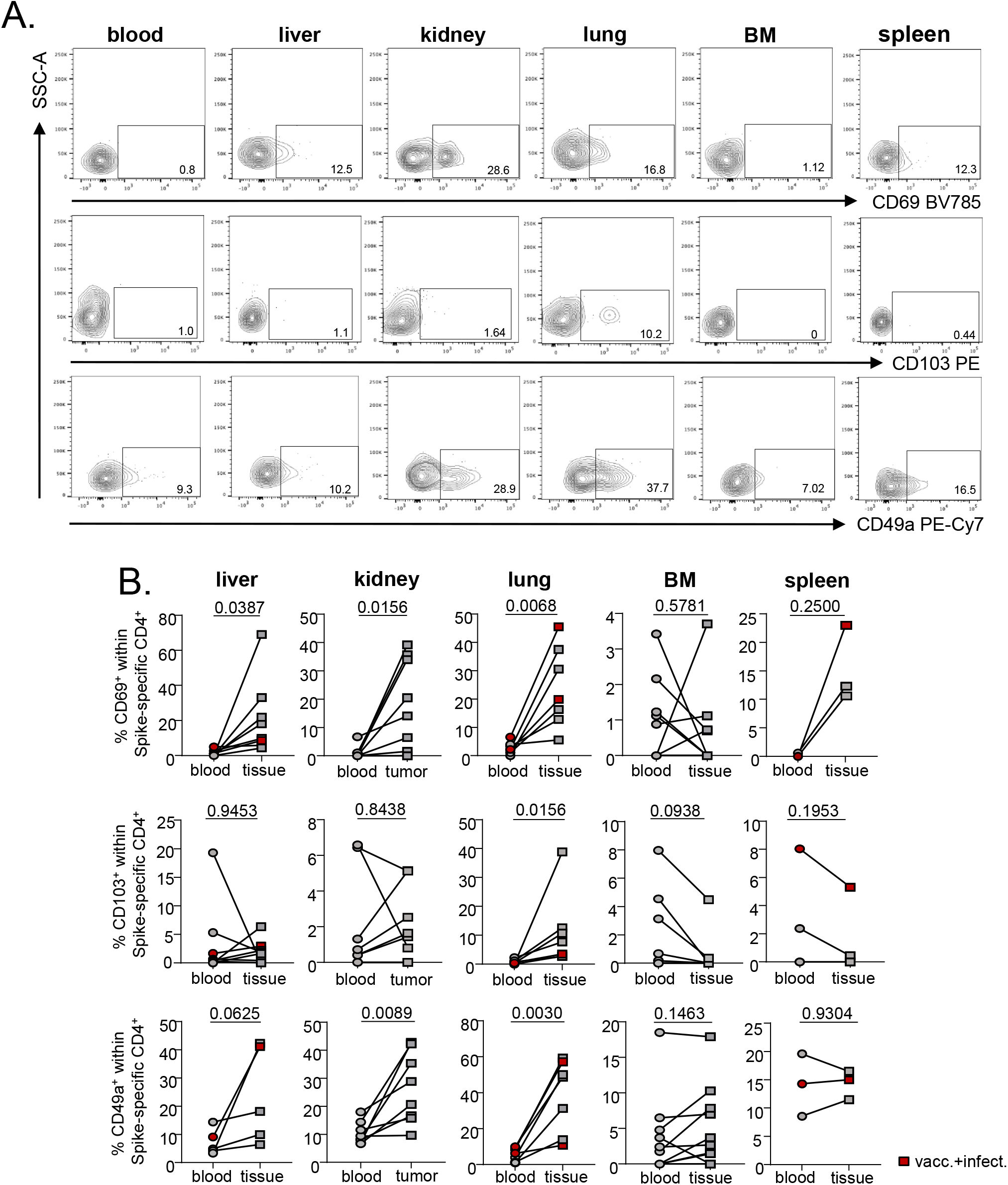
Tissue adaptation signatures of vaccine-specific CD4^+^ T cells. (A) Exemplary plots and (B) summary for FACS-based identification of the tissue residency/retention-associated molecules CD69, CD103 and CD49a amongst vaccine-specific CD4^+^ T cells in the indicated specimen types. Liver: n=8, paired t test for CD69 and Wilcoxon test for CD103/CD49a; kidney: n=8, Wilcoxon test for CD69/CD103 and paired t test for CD49a; lung: n=7, paired t test for Cd69/CD49a and Wilcoxon test for CD103; BM: n=10, Wilcoxon test for CD69/CD103 and paired t test for CD49a; spleen: n=3, Wilcoxon test for CD69 and paired t test for CD103/CD49a. Red symbols identify vaccinated individuals with a history of SARS-CoV2 infection.

Taken together, vaccine-induced CD4^+^ T cells showed a distinct memory/residency signature within non-lymphoid organs in comparison to blood that was in part differentially regulated in their lymphoid counterparts.

### Polyfunctionality as a distinct feature of Spike-specific Th cells from organs

To test the hypothesis that tissue-derived, vaccine-specific CD4^+^ T cells show enhanced functionality as compared to those detected in blood, cytokine production was assessed by FACS (Supplemental Figure 4). No significant differences were detected for interleukin-2 (IL-2), interleukin-4 (IL-4) and interferon gamma (IFNγ) producing cells with the exception of lung showing elevated frequencies of IFNγ^+^ T cells in tissue over blood (Figure 4A and Supplemental Figure 5A). Interestingly, frequencies of IL-2- and IFNγ positive cells correlated between non-lymphoid tissues and paired blood samples (Figure 4B and C), which could not be verified for lymphoid tissues (Supplemental Figure 5B and C). Interestingly, we determined polyfunctionality as a key characteristic separating blood-from non-lymphoid organ-derived T cells. Non-lymphoid organs were enriched for cells expressing two or three cytokines at a time (Figure 4D and E) with only the latter aspect being equally observed for lymphoid organ-derived cells (Supplemental Figure 5D and E). Further investigation revealed an enrichment of specific IL-2-, but not IFNγ producing Th cells within the CD69^-^ subpopulation. On the contrary, IL-2 producers were enriched in the CD49a^+^ Th subset (Figure 4F and G, Supplemental Figure 5F). These analyses were solely conducted for non-lymphoid organs given the paucity of CD69 and/or CD49a expression in lymphoid organs as demonstrated in Figure 3A and B. As a résumé, organ-derived T cells show functional superiority to their blood-derived counterparts mirrored by increased quantities of multipotent cells.

**Figure 4.**
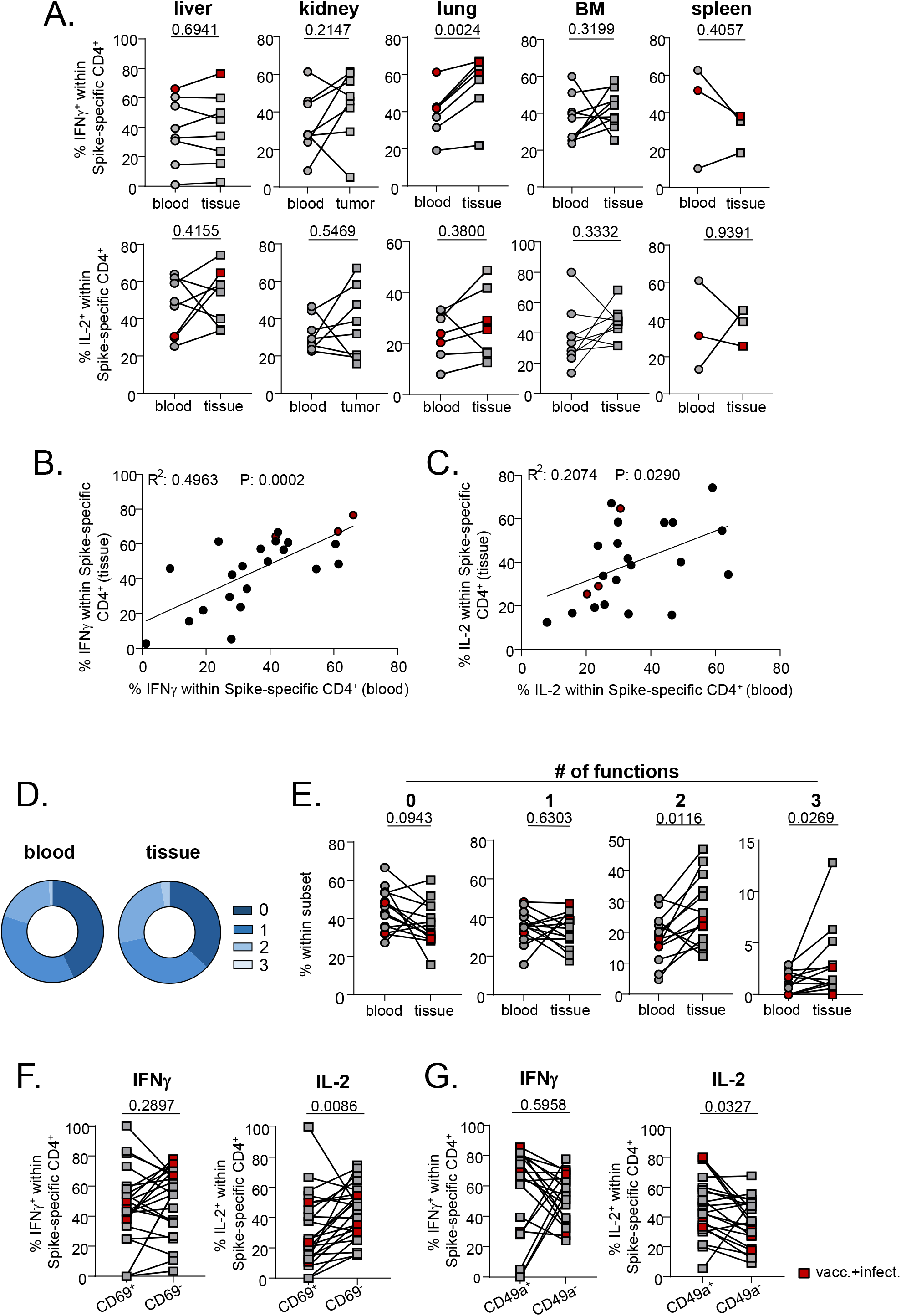
Enhanced polyfunctionality as a feature of specific organ-derived Th cells. Cytokine expression was assessed in Spike-specific Th cells intracellularly by FACS. (A) depicts frequencies of IFNγ or IL-2 positive cells amongst the indicated paired samples. Liver: n=8, paired t test; kidney: n=8, paired t test for IFNγ and Wilcoxon test for IL-2; lung: n=7, paired t test; BM: n=10, paired t test; spleen: n=3, paired t test. (B) Simple linear regression analysis between frequencies of specific IFNγ or (C) IL-2 expressing Th cells from non-lymphoid tissues vs. paired blood. (D) Mean frequencies and (E) paired analyses of Spike-specific polyfunctional Th cells expressing 3, 2, 1 or 0 of the cytokines IFNγ, IL-2 and/or IL-4 at a time. Statistically significant differences were tested with paired t test (0-2 cytokines) or with Wilcoxon test (3 cytokines). (F) Differential IFNγ or IL-2 expression in Spike-specific Th cells from non-lymphoid organs after pre-gating on CD69^-^ or CD49a^-^ expressing or non-expressing subsets, respectively. Liver: n=8; kidney: n=8; lung: n=7. Statistically significant differences were tested with the paired t test (IL-2) or with Wilcoxon test (IFNγ). For D-F, only tissue samples from non-lymphoid organs were included. Red symbols identify vaccinated individuals with a history of SARS-CoV2 infection.

### Vaccine-specific CD8^+^ T cells in non-lymphoid and lymphoid tissues

Along with their CD4^+^ counterparts, antigen-specific CD8^+^ T cells were identified within the same samples according to CD137 and IFNγ co-expression, as recently shown (*4, 16*) with the gating strategy depicted in Supplemental Figure 1. CD8 responder rates, particularly in peripheral blood, were consistently lower (Supplemental Figure 6A). We observed a similar pattern as for CD4 responses in that Spike-specific CD8^+^ T cells were principally detectable in most organ types with the exception of peri-tumor kidney tissue and tonsil where response criteria were consistently not met (Supplemental Figure 6B). Within a limited set of paired samples, no significant elevation of frequencies was observed in tissues over blood (Supplemental Figure 6C). With respect to CD45RO and CD62L expression, no clear sample-type specific pattern was evident for Spike-specific CD8^+^ T cells with the exception of liver tissue showing an enrichment of memory-type T cells over blood (Supplemental Figure 7). Furthermore, although statistical analyses were not adequate due to limited sample size, non-lymphoid organs tended to show selective enrichments of specific CD69^+^, CD103^+^ and/or CD49a^+^ CD8^+^ T cells compared to blood (Supplemental Figure 7).

### SARS-CoV-2-vaccination induced memory B cells are detectable in tissues

Within a limited set of kidney tumor, bone marrow, spleen, tonsil and peripheral blood specimens (Figure 5A), we sought to characterize quantities and phenotype of vaccine-specific B cells. Detection was based on fluorescent double-labelling of specific cells with recombinant full Spike protein coupled to APC, paired with recombinant Spike protein receptor binding domain (RBD-) coupled to AF488 (*17*). The gating strategy is depicted in Supplemental Figure 8. Importantly, Spike-specific B cells were detectable in all organ and blood samples with frequencies typically ranging between 0.1-0.01% within the CD19^+^ compartment. Overall, portions of isotype class switched IgD^-^CD27^+^ memory cells constituted the majority of specific B cells within lymphoid organs and blood, but not in kidney. Expression of CD69 as a marker for tissue retention (*18*) was confined to minor portions of Spike-specific B cells of most specimens with few individual exceptions (Figure 5B and C). Given the small sample size, statistical analyses were not conducted.

**Figure 5.**
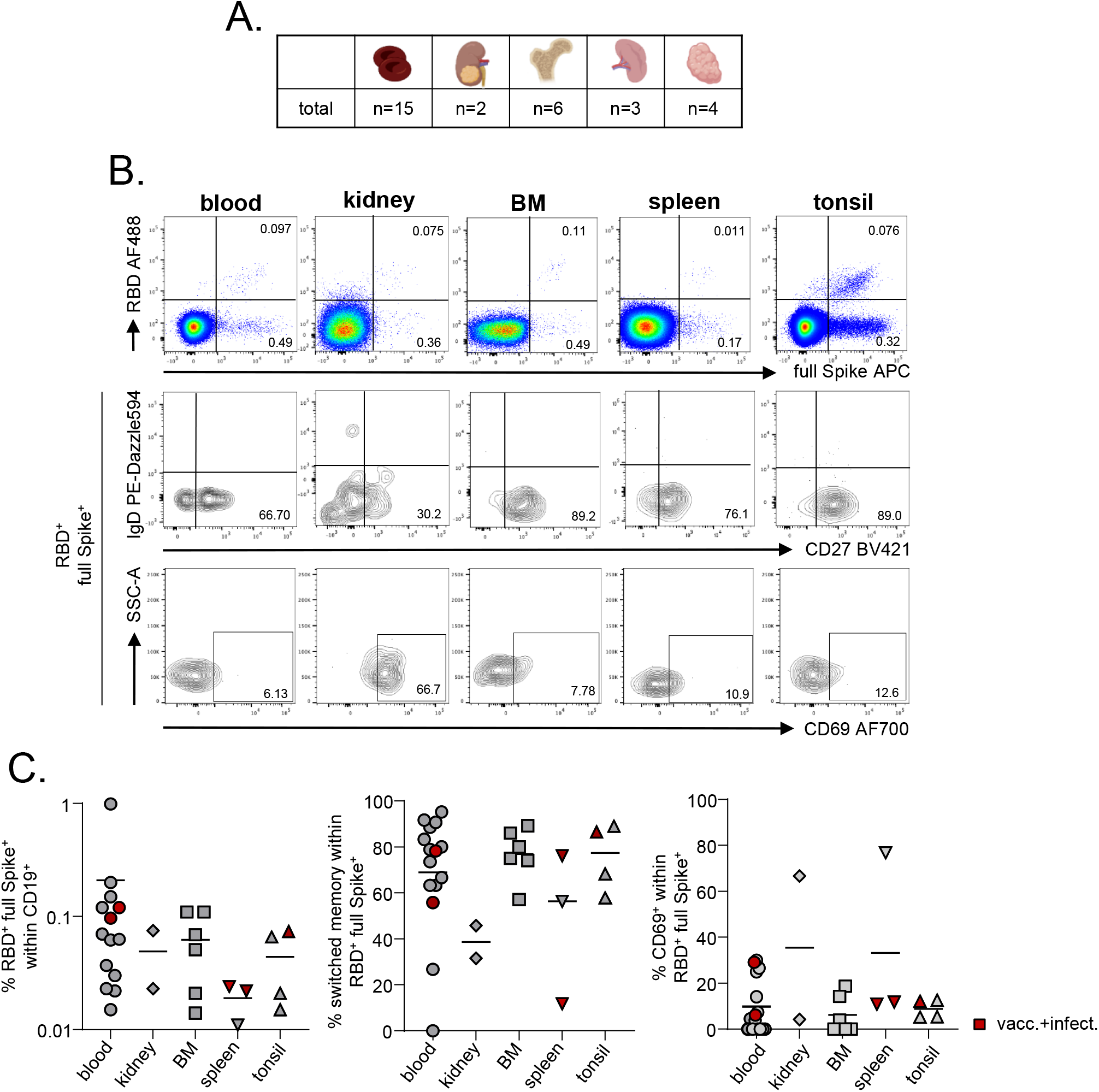
Quantification of SARS-CoV2-vaccine-specific B cells in tissues. (A) Summary of specimens included for specific B cell analysis. (B) Exemplary plots illustrating frequencies of Spike-RBD-specific B cells in the indicated organs (upper panel), their memory phenotype according to CD27 and IgD expression (middle panel) and CD69 expression (lower panel). (C) summarizes data for the indicated specimen types. Red symbols identify vaccinated individuals with a history of SARS-CoV2 infection. Given the small sample size, statistical analyses were not conducted.

### Single cell transcriptomics of vaccine-specific CD4^+^ Th cells

Activation marker-based isolation of SARS-CoV2-specific T cells after antigen-specific stimulation, followed by single-cell RNA sequencing (scRNA-Seq), has been already employed to obtain a deeper understanding of virus-specific, tissue-resident CD4^+^ T cells(*19*) and was applied to characterize specific CD4^+^ and CD8^+^ T cell responses in blood of COVID patients with mild vs. severe disease (*20, 21*) or after SARS-CoV2 mRNA vaccination (*4*). Using this approach, specific CD137^+^CD40L^+^ cells from peripheral blood (n=4), liver (n=4), lung (n=5) and bone marrow (n=3) were FACS sorted to purities >97 % after peptide stimulation (Figure 6A, Supplemental Figure 1), followed by transcriptome assessment (Figure 6B). Blood and liver samples were derived from the same four patients whereas all other specimens were from different individuals. After quality filtering (Supplemental Figure 9), unsupervised clustering of 1.985 Spike-specific Th cells yielded 3 clusters visualized as Uniform Manifold Approximation and Projection (UMAP) (Figure 6C) for which pathway enrichment analyses were conducted (Supplemental Figure 10). Cluster 0 was characterized by the upregulation of cytokine signaling related pathways (therefore in the following termed “cytokine signaling”), whereas cluster 1 showed an enrichment of ribosomal biogenesis-related genes (termed “ribosomal biogenesis”). Cluster 2, showing the most pronounced separation in UMAP, was enriched for developmental-, cell-adhesion related- and T cell activation pathways with the long non-coding RNA *NEAT1* as the most upregulated gene within this cluster (herein termed “NEAT1”).

**Figure 6.**
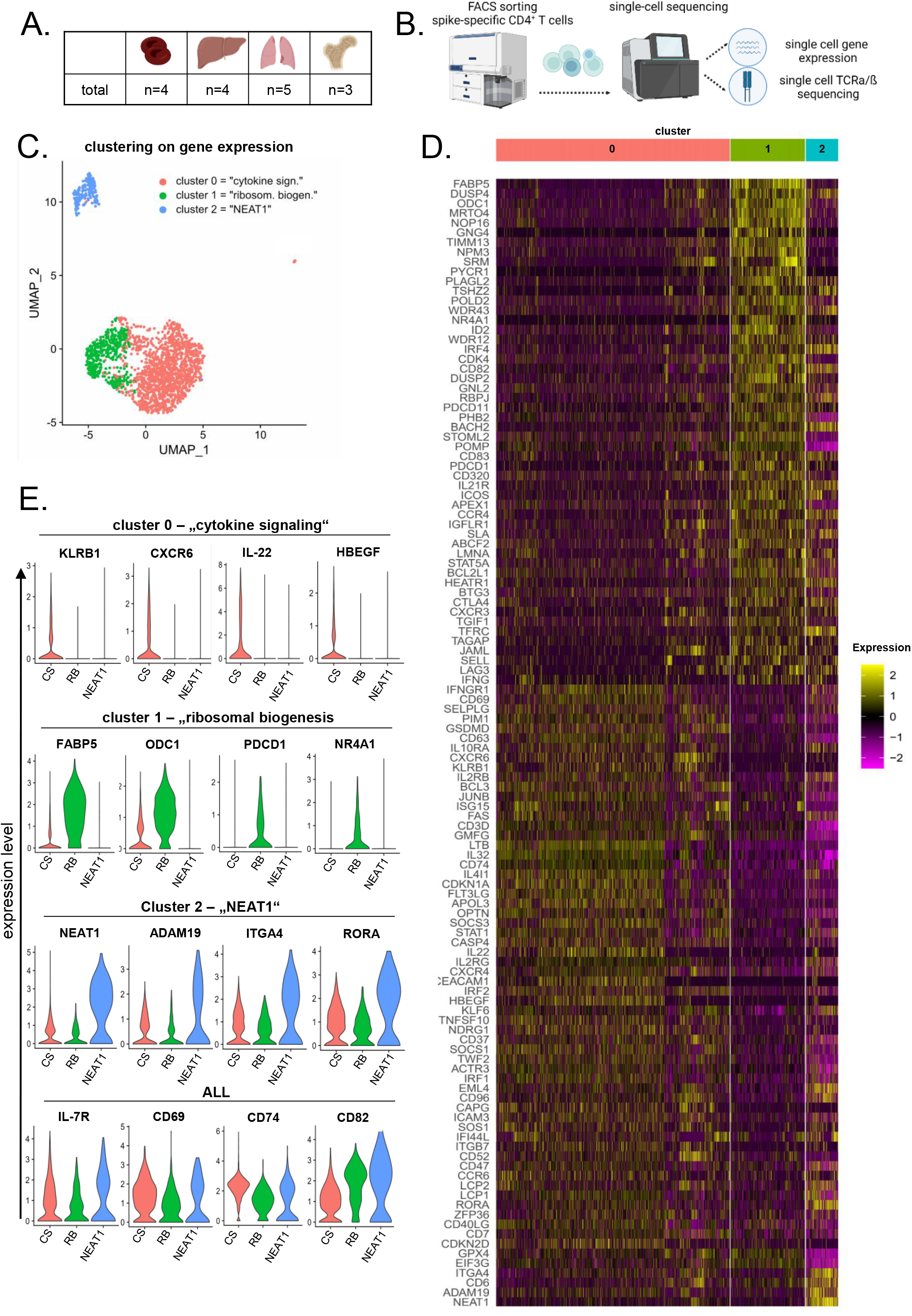
scRNASeq analysis of Spike-specific Th cells from organs and blood. (A) Summary of specimens included and (B) workflow for transcriptome analysis of Spike-specific CD4^+^ Th cells. (C) Unsupervised clustering based of transcriptomes derived from n=1.985 cells identified three major populations when visualized by UMAP. (D) Heatmap showing expression patterns of selected genes characteristic for clusters 0, 1 and 2. (E) Violin plots displaying selection of genes that are differentially regulated in cluster 0 (first panel), 1 (second panel) and 2 (third panel) and those that are similarly regulated over all clusters (panel 4).

Tissue homing and residency related genes including *KLRB1* (encoding CD161) and the chemokine receptor *CXCR6* (*22-24*) were solely upregulated in cluster 0. This cluster showed further transcript enrichments for proinflammatory mediators including IL-22, as well as for heparin-binding EGF (HBEGF), a marker suggested to shape antigen-specific CD4^+^ T cell responses and constraining Th17 differentiation (*25*). In contrast, upregulated genes involved in various metabolic pathways were observed for cluster 1. For instance, a pronounced upregulation of *FABP5* and *ODC1* both involved in lipid metabolism of tissue-resident lymphocytes (*26, 27*) was detected. This cluster was also characterized by induction of the tissue-residency transcript *PDCD1*, encoding PD-1, which has been suggested as a feature of murine tissue-resident brain cells independent of antigen stimulation (*28, 29*), and the transcription factor *NRF4A1*, important for controlling tissue retention (*30*). On the contrary, genes encoding products likely to be involved in tissue-resident cell activation, 11 migration, or retention including *ADAM19* (*31*), the integrin *ITGA4*, and the T cell lineage regulator *RORA* (*32*) was identified for cluster 2. Several transcripts involved in memory differentiation/tissue retention or activation were similarly expressed across clusters, including *IL7R, CD69, CD74 and CD82* (Figure 6D and E).

Surprisingly, employing UMAP, cells from different tissues were not selectively associated with, but evenly distributed over all three clusters (Figure 7A). This observation is in line with the fact that transcripts for a set of typical tissue-related (e.g. *ITGAE, ZNF683, CXCR6*) or circulation/migration-related genes (e.g. *S1PR1, SELL*) were rather cluster- than organ-specific (Figure 7B). RNA velocity for the various tissues did reveal some transcriptional dynamics within clusters, but not across clusters, indicating that the cluster identity itself was likely static (Figure 7C). This observation generally holds for the individual organs, with lung showing a slightly different trajectory and bone marrow suffering from a lower number of cells. These observed effects could be caused by organ-specific differences, but may also be due to the robustness of RNA velocity analyses being related to the density of cells in a region of the UMAP.

**Figure 7.**
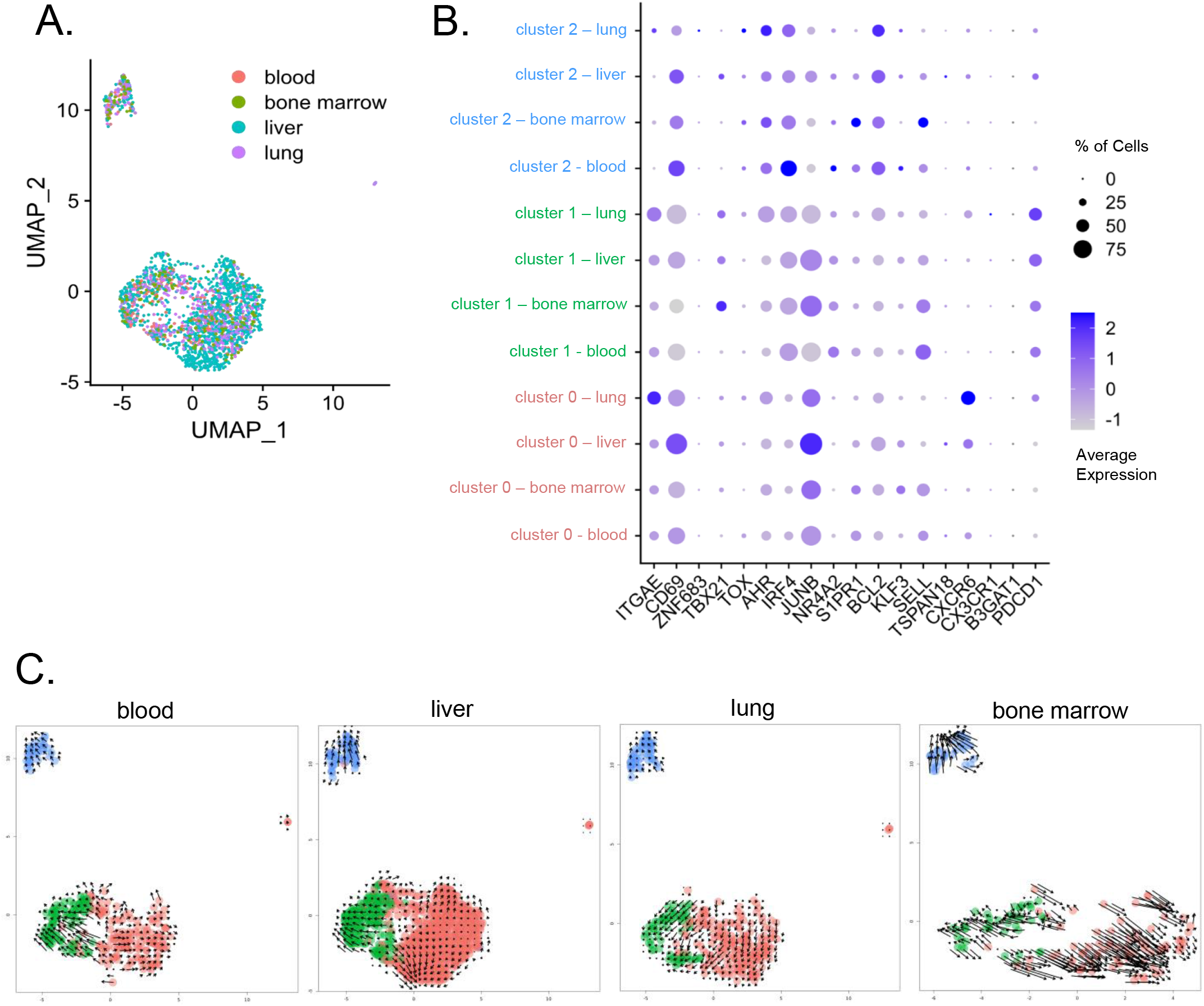
Cell clustering is not primarily driven by tissue-specific features. (A) UMAP plot as in Figure 6C with overlay of specimen origins, respectively. (B) Expression of selected tissue residency/retention-associated or non-associated genes in cells derived from distinct cluster/tissue combinations. Expression values are shown as Z-scores. (C) Grid representation of RNA velocities for the various tissues calculated using velocyto. Datasets were split according to tissue prior to velocity calculation, and cells are color coded by cluster.

### T cell receptor analysis of SARS-CoV2-vaccine-specific T cells

For 1.875 out of 1.985 sequenced cells, a T cell receptor (TCR) clonotype could be obtained. As the sequenced T cells should have a similar antigen specificity, we were interested in the degree of overlap of complementarity-determining region (CDR) 3 sequences between different individuals, as well as different tissues from the same individual. As expected, the highest number of overlapping CDR3 sequences was observed between blood and liver within the same individual (blood/liver #1-4), indicating that the clonal repertoire is in part shared between both tissues. The degree of overlap was less pronounced between different individuals and did not appear to correlate with the tissue of origin. We thus deduce that, again, at least part of the SARS-CoV-2-specific TCR repertoire is not tissue-specific (Figure 8A). The ten most abundant clonotypes within each sample covered a percentage of cells that was correlated with the total number of cells, with all samples roughly showing the same degree of dependency, indicating that the heterogeneity of clonotypes is approximately similar in all samples regardless of tissue (Spearman’s ρ of cell number and percent covered by top clonotypes 0.91, p=1.03*10^−5^; Figure 8B). We next asked whether the clonotype had an impact of the transcriptomic identity of a cell. To this end, we identified all clonotypes with at least four cells, leading to a list of nine different clonotypes. Highlighting the position of cells with a given clonotype in the UMAP projection revealed a strikingly close clustering in most cases, indicating that the gene expression profiles of these cells were much more similar than would be expected by pure chance (Figure 8C). Finally, in order to test which metadata are most influential in driving gene expression, we tested transcriptomic correlation between all cells, and modeled the resulting Spearman correlation coefficients by the following parameters: same or different clonotype/cluster/tissue/donor. Using Tukey’s HSD test, we determined the impact of same versus different metadata for each individual parameter, as well as any interaction of parameters, and found that sharing the same clonotype (or the same CDR3 sequence with a change in Spearman’s Rho of 0.074) had the largest positive effect on correlation of gene expression, followed by the cluster a cell was assigned to. In contrast, the tissue of origin had a smaller effect, and only a very small proportion of the correlation was driven by cells coming from the same donor (Figure 8D). Thus, the clonotype, i.e. the T cell receptor sequence, was the best predictor of cells sharing similar transcriptomes.

**Figure 8.**
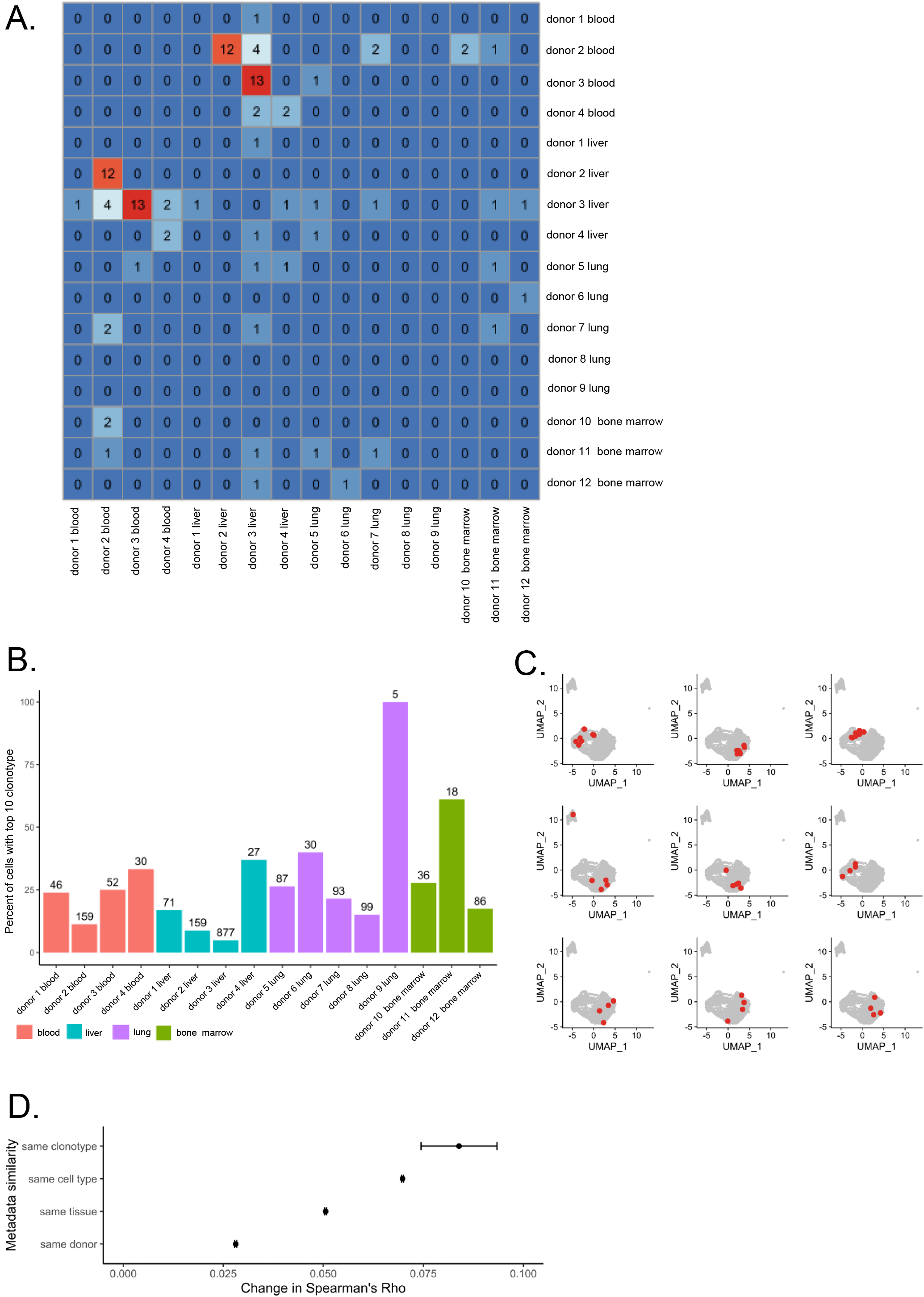
Shared TCR clonotypes between tissues. (A) Heatmap depicting the overlap in absolute numbers of CDR3 sequences in different samples. (B) Percentage of cells with ≥1 out of the ten most frequent clonotypes per sample, coloured by organ. Total cell numbers with known clonotype are indicated above the bars. Blood and liver samples from donors 1-4 were paired, whereas samples 5-12 were from different donors. (C) Association ofbclonotypes with gene expression. UMAP plots with cells highlighted in red that have a shared clonotype. Separate graphs for all nine different clonotypes with at least four cells (=inclusion criterium) are shown. (D) Impact of shared vs. different metadata on the cell-cell Spearman correlation coefficient for highly variable genes. Mean change and 95% confidence intervals were obtained using Tukey’s honest significance of differences test, considering all individual variables as well as their interactions.

## Discussion

So far, it remained largely obscure whether and how cellular memory induced by intramuscular vaccination acquires residence in human tissues and how it may adapt to distinct local environments, particularly considering novel mRNA-based vaccines. Although the SARS-CoV2 pandemic has stimulated research on numerous aspects of antiviral protection, the availability of human tissue specimens still constitutes the major limitation for comprehensive assessment of organ-specific immunity. To the best of our knowledge, our study therefore provides pioneering data on tissue distribution, molecular signatures and functional capacity of SARS-CoV2-specific T cells generated after mRNA vaccination. Most importantly, and particularly considering that samples were largely derived from virus-naïve individuals with preconditions, Spike-specific CD4^+^ Th cells were detectable approximately 3-4 months after vaccination in all non-lymphoid and lymphoid organ types analyzed except for tonsil. Besides peripheral blood, highest responder rates were determined for bone marrow, being in accordance with its role as survival niche of T cells specific for systemic pathogens (*12*). Since the aforementioned and a related study (*33*) could not differentiate between vaccination and infection-induced responses due to the live measles, mumps, and rubella (MMR) vaccine employed, there was no compelling indication so far that results could be extrapolated to mRNA vaccines.

Consistent with our recent findings on human kidney-derived bulk T cells (*11*), human renal peri-tumor tissue proved to be lymphopenic as compared to tumor specimens, supporting favorable quantification of vaccine-specific T cells in the latter. Whereas our previous report (*34*) showed similar frequencies of cytotoxic CD8^+^ T cells specific for persistent (Epstein-Barr virus, EBV; cytomegalovirus, CMV) or seasonal (influenza) viruses in kidney and blood, we found enrichment of SARS-CoV2-vaccination induced CD4^+^ Th cells as a characteristic feature within non-lymphoid, as compared to lymphoid organs. Importantly, frequencies of total vaccine-specific Th cells significantly declined with progressing vaccinee age in blood, but not in paired non-lymphoid tissue samples. This observation also applied to specific CD45RO^+^CD62L^-^ Th cells and is in agreement with the concept that maintenance of organ memory shows increased robustness (*35*). Stable age-independent persistence of immunological memory has been demonstrated for CMV- and influenza-specific CD8^+^ tissue-resident memory T (Trm) cells isolated e.g. from human lung tissue (*19*). It needs to be considered, however, that T cells specific for persistent or recurring viruses are established early in life (*36*) and subject to frequent reactivation *in vivo*, whereas longer-term maintenance of mRNA vaccine-induced memory might critically depend on periodic boosters. With respect to their specific organ adaptation, a substantial portion of SARS-Cov2-specific Th cells showed expression of the tissue residency associated molecules CD69 and CD49a, whereas CD103 expression was mainly confined to the lung. This pattern broadly reflects expression characteristics defined for bulk and influenza-specific CD4^+^ Th cells isolated from human lung- and kidney tissue (*11, 37, 38*). Based on our data, a segregation between lymphoid and non-lymphoid tissues was particularly evident for CD49a expression, showing an upregulation over blood only in liver, kidney and lung. Interestingly, data from nasal tissue procured after mRNA vaccination support our observations in regard to specific T cell detection at distal sites in general and low frequencies of CD103^+^ cells in particular (*39*). Predominant, yet limited CD103 expression of lung-derived CD4^+^ and CD8^+^ T cells has also been reported after human SARS-CoV2 infection (*9*). A murine mRNA-based influenza vaccination model suggests that differentiation into CD69^+^CD103^+^ Trm could be enforced by intra-muscular priming, followed by secondary intranasal boost (*40*). Since this strategy is considered promising due to its enhanced protection of mucosal sites (*41*), corresponding phase III clinical studies are currently under way for improved local protection against SARS-CoV2 infection (*42*). Adequate comparisons between our findings and other studies on CD69 expression are limited by the fact that CD69 is uniformly upregulated by antigenic stimulation, a feature that we circumvent by stably staining cells prior to activation. Therefore, previous estimation of antigen-specific CD69^+^ Th cells from tissues has likely been biased (*9, 43*).

In principle, the concept of tissue memory entails optimized positioning of cells at potential future infection sites in concert with functional specialization (*44*). Amongst those, enhanced polyfunctionality has been determined a distinct feature of tissue-derived lymphocytes as compared to their circulating counterparts (*33, 38, 45*), e.g. correlating with superior viral clearance after experimental influenza vaccination (*46, 47*). It remained unclear whether such functional bias may also apply to SARS-CoV2-specific T cells induced after local vaccination. With respect to single cytokine analysis, only lung-derived Th cells contained significantly higher portions of IFNγ^+^ cells compared to blood, a feature being shared between vaccinated and previously infected individuals. The fact that frequencies of IFNγ and IL-2 positive Th cells (and of the total specific Th population) highly correlated between non-lymphoid tissues and blood indicates that the individual’s potential to mount a vaccination response might be regulated on a systemic level, leading to corresponding magnitudes amongst different sites. At the same time, our cytokine expression data extend the notion that tissue-derived Th cells show functional modifications over their blood-derived counterparts: in accordance with cytokine expression in bulk T cells derived from various human organs (*48*), we found a significant enrichment of Spike-specific polyfunctional Th cells (that is, cells co-expressing IFNγ, IL-2 and IL-4 at a time) both in non-lymphoid and lymphoid tissues. It remains to be addressed whether such superior functional programming depends on priming proximal to the respective tissue sites, as is suggestive by similar features of SARS-CoV2 infection induced Th cells isolated from of lung-associated lymph node and lung tissue (*9*). Priming distal to the inoculation site would require distribution of vaccine-encoded antigens, an aspect being addressed in experimental models. Importantly, for mRNA encoded model antigens, expression has been demonstrated in liver and lung (*14*) whereas vector vaccine-encoded antigens were detectable for up to weeks in multiple distant non-lymphoid and lymphoid organs (*15*). It is therefore principally conceivable that *in vivo* distribution of vaccination antigens to distant priming or boosting sites contributes to customized tissue memory differentiation, a hypothesis being principally supported by identification of vaccine-specific memory B and cells CD8^+^ T at multiple sites, including distant tissues. Overall, CD8 responder rates were particularly lower for blood, bone marrow, liver and kidney as compared to CD4^+^ Th cells. Whereas common tissue-specific patterns were not as clear cut as for CD4^+^ Th cells, increased expression of Trm related molecules was most pronounced for specific CD8^+^ T cells from liver and lung. Since, as indicated earlier, these organs are most likely to express vaccine-encoded antigens (*14*), experimental models are needed to experimentally address their role in local priming/boosting in a similar manner as has been elegantly demonstrated for B cell responses in draining lymph nodes (*49*).

Opposed to our initial expectations, we did not identify pronounced organ-specific transcriptome signatures of vaccine-specific CD4^+^ T cells. Instead, we observed a robust separation into three clusters that was largely independent of tissue origin. Pathway enrichment analyses revealed a differentiation into an inter-active (cytokine signaling), translationally active (ribosomal biogenesis) and metabolically active (NEAT1) cluster suggesting functional rather than tissue-specific adaptations of Trm showing mostly transcriptional dynamics within clusters as reflected by RNA velocity analysis. Interestingly, these data were further supported by TCR analysis, revealing that identical clonotypes are largely confined to the same cluster, indicating related functional programs associated with a given clone, even across (paired) tissue samples. Our finding that sharing the same clonotype had the largest positive effect on correlation of gene expression could be interpreted in that the transcriptomic landscape, and thus the cell state and cluster annotation, is determined by its parent cell from which it inherited the TCR sequence – and not necessarily by tissue origin.

In general, our results on the TCR landscape provide valuable information to estimate the breadth of the vaccination response: on average around 20 % of cells belong to one of the ten most frequent clonotypes within a sample, indicating a quite oligoclonal repertoire, considering the large size of the viral Spike protein.

Characteristic murine virus-specific and human bulk transcriptional T cell signatures have been identified in tissues, pointing towards distinct programs at barrier- as compared to lymphoid sites and blood, including clonotype distribution (*50, 51*). Whereas mechanisms of Trm ontogeny remain incompletely addressed, two main hypotheses have been postulated (*52*): Whereas the local divergence model states that mainly soluble factors within a given tissue drive memory T cells towards a Trm phenotype (*50, 53*), the systemic residence memory differentiation model hypothesizes that T cells are transcriptionally marked based on their variable or identical TCR and therefore skewed toward either a Trm or circulating memory T cell (*54-56*). Our results are in line with the latter, indicating that the TCR sequence is a strong determinant of the transcriptomic state in tissue-derived, vaccine-specific CD4^+^ T cells. Still, it needs to be clarified, when and where antigen-specific immune cells acquire exactly tissue-residency, and whether this is a process underlying a certain kinetic further dependent from intensity and site of antigen encounter, TCR affinity strength as well as from activation and metabolic requirements of responding immune cells (*40*).

Overall, the idea that tissue-resident T cells comprise an “inert” population that does not recirculate has recently been challenged by demonstrating human bone marrow Trm to be recruited into the blood upon MMR re-vaccination (30). Accordingly, human skin Trm cells shuttling between tissue and blood are characterized by similar transcriptional programs (43). These studies principally reflect the emergence of a more dynamic concept of tissue memory and associated molecular patterns as is being currently discussed (42, 44); it yet has to be more substantiated with respect to vaccination-specific responses where the impact of a local re-challenge (*40*) might be critical.

Our study has several limitations. Given that it relied on human surgery specimens, procurement of bona fide “healthy” human tissue is not feasible. As a consequence, we cannot completely rule out distinct effects of primary diseases of our organ donors on quantity and quality of tissue-derived lymphocytes. However, our comparative analysis on T cell responses in tumor- vs. non-tumor patients, in line with comprehensive data on humoral and cellular immunity in solid cancer patients (*57*), suggest that the potential impact of preconditions or treatment on vaccine-induced responses is likely small. Based on our previous experience, peri-tumor versus tumor-derived renal bulk T cells exhibited only subtle functional differences with no fundamental changes as compared to blood (*11*). Altogether, these observations argue in favor of functional robustness of virus-specific T cells despite potential immunosuppressive effects of the tumor environment (*34*). Furthermore, we cannot estimate the exact impact of the stimulation approach on scRNA-sequencing results. Activation-induced-marker (AIM)-based approaches for sorting of human antigen-specific T cells as a prerequisite for RNASeq analysis have been employed in multiple settings, including studies focusing on tissue residency: Farber et al. identified both virus- and tissue-dependent signatures of CMV and influenza-specific human T cells from blood and tissues of deceased organ donors after 24h of in vitro stimulation (*19*). Both in the context of SARS-CoV2 infection and vaccination, AIM-based strategies for isolation of virus-specific cells, followed by scRNASeq, were used to identify patient group-specific transcriptional signatures (*4, 20, 21*). On that background, we have no indication to assume that our cell sorting strategy principally prevents identification of distinct transcriptomic signatures, but potentially excludes molecules equally induced across samples by activation, such as CD69 (*21*). Given that human T cells earliest divide after 42-65 hours (*58*), it seems further unlikely that our results are biased by selective expansion of clones within the 16 h stimulation period. Activation-dependent transcriptional changes could theoretically be minimized by use of multimer-based cell purification, albeit restricting ensuing analyses by the concomitant selection of immunodominant peptides as e.g. recently identified in (*59*), therefore not representing a favorable strategy. Although we could identify vaccine-specific B cells amongst multiple tissues, we were not able to similarly assess a possible dichotomy between non-lymphoid and lymphoid organs due to limited sample availability. Possible confounders with respect to differential systemic effects of primary disease or previous medication are excluded by our strategy to pairwise analyze blood and tissue specimens, assuming that both compartments are similarly affected by systemic preconditions. The same applies to the time passed since last vaccination. An impact of such bias could be further excluded by the fact that frequencies and functions of specific Th cells in blood and tissue remained stable over time since last vaccination, being in accordance with other studies on long-term T cell maintenance after SARS-CoV2-vaccination (*60-62*). Based on the current recommendations for 6-month booster vaccination intervals particularly for aged individuals or those with comorbidities, recruitment of patients for analyzing tissue memory at later time points is limited. The same applies to the inclusion of more vaccinated, virus-naïve individuals due to the progressing Omicron variant dissemination. A larger cohort would better compensate for the inter-individual variation and increasing cell numbers for transcriptome analysis would allow a better representation of the cellular repertoire to a comparably large antigen as the SARS-CoV2 Spike protein.

In summary, we reveal here key features of mRNA vaccination-induced CD4^+^ Th lymphocytes with respect to their distribution across the human body, memory differentiation, age association and functional adaptation. Moreover, our scRNA-sequencing data suggest that these Spike-specific CD4^+^ T cells are maintained as defined subsets across lymphoid and non-lymphoid tissues pointing towards functional rather than organ-specific adaptations. These data suggest a heterogenous plasticity of vaccine-specific Th cells in human tissues which might be of further relevance for the development of efficient vaccination strategies in the future.

## Material and Methods

### Patients

Macroscopic portions of tumor- and/or most distant peri-tumor tissue as well as paired peripheral blood and serum samples were collected between October 2021 and October 2022 from patients diagnosed with a renal, liver or lung tumor. Only few patients with distant metastases and/or on chemotherapy (for less than 8 weeks before/after vaccinations or analysis) were included. Patients had no additional inflammatory diseases and did not receive immunosuppressive medication. Bone marrow was collected during spine surgery, and tonsils and spleens were collected following tonsillectomy and splenectomy. All patients were vaccinated against SARS-CoV2 according to the national vaccination program and completed the two- or three-dose vaccination protocol with BNT162b2 (“Comirnaty”, BioNTech/Pfizer, 30μg/dose) or with mRNA-1273 (“Spikevax”, Moderna/NIAID, 100μg/dose). The interval between the shortest and the longest time point after last vaccination ranged from 19 to 265 days. Patient demographics, including time since last vaccination, are summarized in Table 1.

### Study Approval

The study was approved by the local Ethics Committee of the Charité-Universitätsmedizin Berlin (EA4/066/19, EA1/353/16, EA4/115/21) and University Hospital Leipzig (322/17-ek, 237/22-ek) and was conducted in compliance with the declarations of Helsinki and Istanbul. All patients provided written informed consent.

### Sample processing

Serum samples were stored at −80°C. Resected tissues were immediately processed. To obtain single cell suspensions, tissue was dissected into small pieces. Digestion medium was added, consisting of RPMI 1640 medium (Corning, Falcon, Kaiserslautern, Germany) supplemented with 0.3 mg/ml glutamine, 10% FCS (Gibco, Thermo Fisher, Darmstadt, Germany), 1% P/S (Sigma Aldrich, Merck, Darmstadt, Germany), 1 mg/ml Collagenase II (Gibco) and 10 U/ml DNAse I (Sigma Aldrich, Merck, Darmstadt, Germany). Samples were incubated for 45 min at 37°C while shaking. Reaction was stopped with medium without enzymes and cells were passed through a 100 μm cell strainer (Corning). Thereafter, mononuclear cells (MNCs) were isolated with Leuko-Human Separating Solution (Genaxxon, Ulm, Germany) by density gradient centrifugation and immediately cryopreserved. The latter two steps equally applied to peripheral blood mononuclear cells (PBMC).

### Assessment of humoral immunity

Previous or current SARS-CoV2 infection was assessed based on medical history and SARS-CoV2 nucleoprotein specific ELISA (Euroimmun). SARS-CoV-2 S1 domain specific IgG was determined by ELISA (QuantiVac, Euroimmun). Serum samples with OD ratios of ≥1.1 (nucleoprotein-specific IgG) or ≥35.2 BAU/ml (Spike-specific IgG) were considered positive according to the manufacturer’s guidelines. OD ratios were calculated based on the ratio of the OD of the respective sample over the OD of the calibrator provided with the ELISA kit.

### Assessment of SARS-CoV2 vaccine-specific B and T cells

Mononuclear cells (MNCs) were thawed and washed twice in prewarmed RPMI 1640 medium (containing 0.3 mg/ml glutamine, 100 U/ml penicillin, 0.1 mg/ml streptomycin, 20% FCS, and 25 U/ml benzonase (Santa Cruz Biotechnology Inc.). For identification of vaccine-specific T cells, 3-5×10^6^ PBMC were stained with CD69 BV785 (FN50, Biolegend) for 20 min at RT to identify *ex vivo* expressing cells. CD69 staining was stable over the following stimulation period as demonstrated earlier (*11*) and depicted in Supplemental Figure 1B. Thereafter, cells were rested for 2h at 37°C and stimulated or not for 16 h with overlapping peptide 15-mers covering the complete SARS-CoV2 Spike protein (alpha-variant) at a final concentration of 0.5 μg/ml per peptide (JPT, Berlin, Germany). Brefeldin A (Sigma-Aldrich, St. Louis, Missouri, USA) was added after 2 h. T cells were identified as CD3^+^CD19^-^CD14^-^ live (“dump^-^”) single lymphocytes. Vaccine-specific CD4^+^ T helper cells were identified based on CD137 and CD40L, whereas specific CD8^+^ T cells were detected based on CD137 and IFNγ co-expression with the gating strategy depicted in Supplementary Figure 1A).

A response was defined as positive when stimulated cultures contained at least twofold higher frequencies of CD137^+^CD154^+^ CD4^+^ T cells or CD137^+^IFNγ^+^ CD8^+^ T cells as compared to the respective unstimulated control with at least twenty events, as reported earlier (*4, 16*). For assessment of polyfunctionality, samples with at least forty CD137^+^CD40L^+^ cells were included. For surface labelling, antibodies listed in Supplemental Table 1 were used. After surface staining, cells were fixed with FACS Lysing Solution (BD), followed by permeabilization in FACS Perm II Solution (BD) and stained intracellularly with antibodies as summarized in Supplemental Table 1.

B cells were detected within 5-10×10^6^ MNCs by flow cytometry and gated as CD19^+^CD3^-^ CD14^-^CD56^-^ live (“dump^-^”) single lymphocytes. SARS-CoV2-specific B cells were identified as shown before (*17*) by double staining with recombinant receptor binding domain (RBD) protein (alpha-variant, RnD Systems, Minneapolis, MN, USA) coupled to AlexaFluor488 and recombinant full Spike protein coupled to biotin (alpha-variant, RnD Systems, Minneapolis, MN, USA), with the latter detected by streptavidin-APC (Biolegend, San Diego, CA, USA).The gating strategy is depicted in Supplemental Figure 7. For further flow cytometric surface marker expression analysis, antibodies depicted in Supplemental Table 2 were used. Data was acquired using a BD FACS Fortessa X20 with DIVA software V8.0.7.

### FACS data analysis and statistics

FACS data analysis was conducted with FlowJo 10 (BD). Frequencies of Spike-specific T cells were background- (=unstimulated control) substracted. Co-expression of cytokines was quantified by Boolean gating in Flowjo. Statistical analysis and graph preparation were performed in GraphPad Prism 8 (GraphPad, La Jolla, CA, USA). Data distribution was assessed using the Kolmogorov-Smirnov test. Depending on normal distribution or not, ANOVA (with Holm-Sidak’s post-hoc) or Kruskal-Wallis test (with Dunn post-hoc) were chosen for multiple comparisons. For two-group comparisons, unpaired t test or Mann-Whitney test were used. The relationship between two variables was examined by simple linear regression analysis. For analysis of contingency tables, *Fisher’s exact* test was applied. In all tests, a value of p<0.05 was considered significant.

### Enrichment of vaccine-specific CD4^+^ T cells and single cell RNASeq analysis

For single cell transcriptome (scRNASeq) analysis, 10^7^ MNCs from peripheral blood, liver, lung and bone marrow were stimulated for 16 h with SARS-CoV2 Spike peptide mix in the presence of anti-CD40 (1 μg/ml, HB14, Miltenyi Biotec) to retain CD154 on the surface of specific CD4^+^ T cells (*63*). Thereafter, antigen-reactive cells were surface stained with anti-CD154 PE (24-31, BioLegend) and magnetically enriched using anti-PE nanobeads (BioLegend) over MACS LS columns (Miltenyi Biotec). Spike-specific CD3^+^CD4^+^DUMP^-^ CD154^+^CD137^+^ cells were further FACS purified in single cell mode to typically >97 % purity (exemplarily depicted in Supplemental Figure 1A) into PBS/BSA buffer containing round shaped PCR tube lids on an Aria Fusion cell sorter (BD). To minimize cell loss related to small numbers of specific T cells, samples were individually spiked with dump^-^CD3^-^CD4^-^CD8^-^ CD56^+^ Natural Killer (NK) cells from the same sample, resulting in a total of 5.000 cells per sort. The cell suspension was loaded into a 10X Chromium Controller using 10X Genomics Chromium Next GEM Single Cell V(D)J Reagent Kit v1.1 (10X Genomics), and the subsequent reverse transcription, complementary DNA (cDNA) amplification and cDNA library preparation was performed according to the manufacturer’s instructions. 5’ gene expression libraries and target enriched libraries (TCR, for human T cells) were quantified by Qubit™ 4.0 Fluorometer (ThermoFisher) and quality was checked using 4200 Tapestation with High Sensitivity DNA kit (Agilent). Libraries were then pooled in a 10:1 ratio (5’ gene expression library: target enriched library). Sequencing of the pooled library was performed in paired-end mode with SP flow cells (2 × 50 cycles kit) using NovaSeq 6000 sequencer (Illumina).

### scRNASeq analysis and statistics

Primary analysis as TCR and 5’ gene expression libraries was performed using Cell Ranger V(D)J 6.1.2 (10X Genomics). Reference build 5.0.0 (VDJ) and GRCh38-2020A (gene expression) were used. Conserved clonotypes within cells from a single donor were identified using Cell Ranger aggr, while downstream analysis of gene expression data was performed on the unaggregated samples. For secondary analysis, Seurat 4.2.1 (*64*) was used. Following pre-filtering, excluding any cell with less than 200 genes expressed or more than 10% mitochondrial reads, as well as any gene expressed in less than 3 cells, gene expression was normalized to 10,000 reads per cell. As sequenced cells were a pool of NK and CD4^+^ T cells, the latter were extracted using a filter defining any cell with at least one read of CD4 or a T cell receptor sequence and no reads of *FCGR3A* as CD4^+^ T cell. Samples were then integrated using Harmony 0.1.0 (*65*). After principal component analysis, nearest neighbor graph calculation and Leiden clustering (*66*) with a resolution of 0.1, four clusters of cells were identified. The smallest cluster showed transcriptional patterns akin to NK cells and was thus deemed a likely contamination. Any further analyses were performed using the three larger clusters (cluster 0, 1, 2). Differentially expressed genes were identified using the FindMarkers and FindAllMarkers functions with a Wilcoxon test. Pathway enrichment analyses were performed using MetaScape (*67*). For RNA velocity analyses, spliced and unspliced count matrices were generated using velocyto CLI (version 0.17.17). Velocity was then calculated and projected onto to UMAP using velocyto.R (version 0.6). For analyses by tissue, the dataset was split prior to velocity calculation. To estimate the skewing of cell state distribution of clonotypes, a Monte Carlo simulation was run for each clonotype to determine the tail probability of observing a distribution that was as skewed or more skewed than the one observed. The p-value across clonotypes was then calculated as their joint distribution.

## Supporting information

Pross et al Supplemental Files

## Data Availability

All cellular data needed to evaluate the conclusions in the paper are present in the paper or the Supplementary Materials. Upon publication, scRNA-Seq data will be available at EGA for non-commercial research, subject to controlled access according to EU and German data protection regulations. This study did not use any unique codes, and all analyses were performed in R and Python using standard protocols from previously published packages as indicated. Requests for materials should be directed to K.K., A.S. or S.L.

## Acknowledgements and funding

KK is supported by grants from the Deutsche Forschungsgemeinschaft (KO-2270/7-1, KO-2270/4-1, KO-2270/8-1), Wilhelm-Sander Stiftung (2022.035.1), Zentrales Innovationsprogramm Mittelstand (KK5463201BA2) and unconditional project funding from Chiesi GmbH. The funder was not involved in the study design, collection, analysis, interpretation of data, the writing of this article or the decision to submit it for publication. ES is enrolled in the Charité Clinician Scientist Program funded by the Charité– Universitätsmedizin Berlin and the Berlin Institute of Health. SL is supported by the German Ministry for Education and Research through the Medical Informatics Initiative (junior research group “Medical Omics”, 01ZZ2001). HS received funding from the Ministry for Science, Research, and Arts of Baden-Württemberg, Germany and the European Commission (HORIZON2020 Project SUPPORT-E, no. 101015756).

## Author contributions

VP, AS, CC and KK designed experiments.

VP, AS, LT, LMLT, JS, AH, KJ, CL, SL performed experiments.

VP, AS, SL, CC, analyzed the data.

VP, AS, SL, CC and KK interpreted the data.

GD, DS, CG, CB, PJ, AM, SE, BK, TM, KB, DL, JK, JB, FR, FF, MH, SW, NL, ES, HS provided tissue-, blood samples and clinical data, analyzed clinical data, consulted and discussed the manuscript.

AS, CC and KK supervised the project.

VP, AS, SL and KK wrote the manuscript. All authors reviewed and approved the final manuscript.

## Abbreviations

CDR3: complementarity-determining region
COVID-19: corona virus disease 2019
MNC: mononuclear cells
MMR: measles, mumps, and rubella
SARS-CoV2: severe acute respiratory syndrome coronavirus type 2
TCR: T cell receptor
Th cell: T helper cell
Trm: tissue resident memory T cell
UMAP: Uniform Manifold Approximation and Projection

