## Supplementary material for "SARS-CoV2 mRNA-vaccination-induced Immunological Memory in Human Non-Lymphoid and Lymphoid Tissues": Pross et al Supplemental Files

### Proß, Sattler, Lukassen et al. Supplemental Figure 1

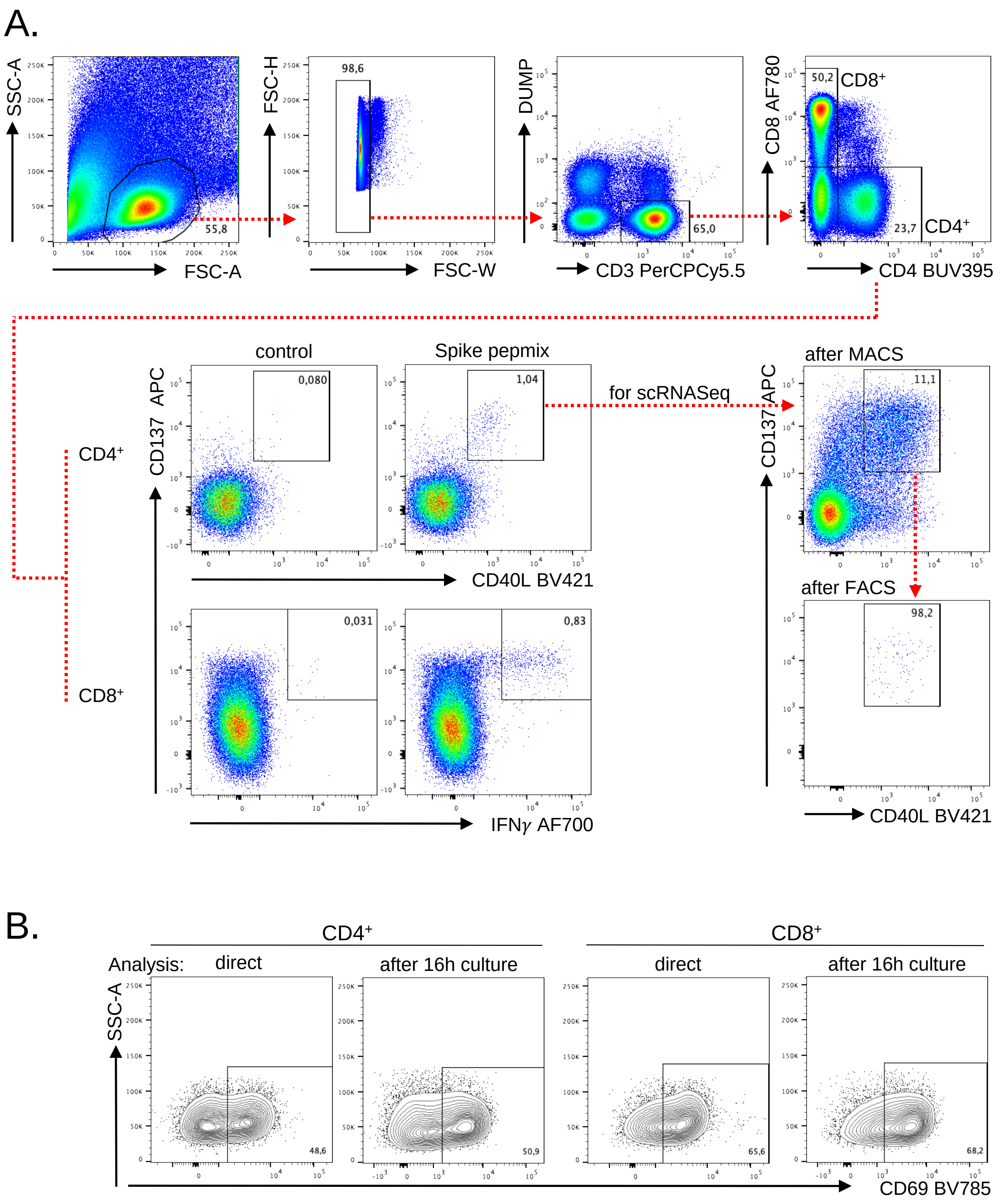


**Supplemental Figure 1. Gating strategies for identification of SARS-CoV-2 vaccine-specific T cells.** (A) Exemplary, liver derived MNCs were stimulated or not with Spike-peptide mix for 16 h as indicated. Live single CD14⁻CD19⁻CD3⁺ specific CD4⁺ Th cells were identified by FACS according to co-expression of CD137 and CD40L. Doublets were excluded based on FSC-W/FSC-H signals. For scRNASeq analysis, CD40L⁺ cells were magnetically enriched, followed by FACS sort of live CD4⁺CD137⁺CD40L⁺ cells to typically >97 % purity. Spike-specific CD8⁺ T cells were detected based on CD137 and IFNγ co-expression. (B) Exemplary, lung-derived MNCs stained with CD69 BV-785 showed no relevant loss of CD69 signal when analyzed by FACS after 16 h of cultivation as compared to direct analysis.

#### Proß, Sattler, Lukassen et al. Supplemental Figure 2

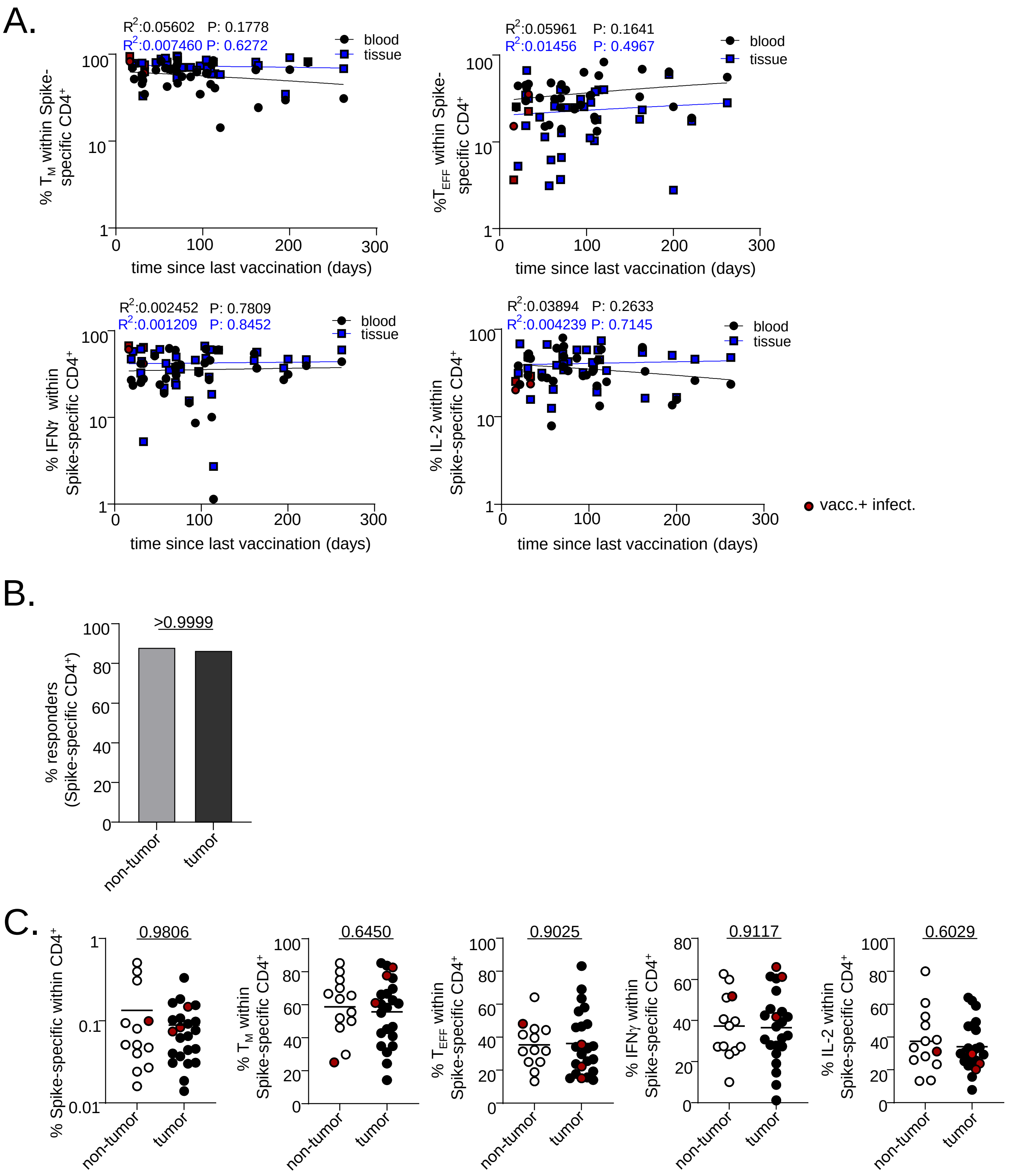


**Supplemental Figure 2. Impact of time since last vaccination or patient precondition on quantity and/or quality of vaccine-specific CD4$^+$ T cells.** (A) Correlation analyses of blood and tissue derived frequencies of Spike-specific memory (upper left), effector (upper right), IFNγ$^+$ (lower left) or IL-2$^+$ (lower right) CD4$^+$ T cells with time since last vaccination. BM: n=10, spleen: n=3, liver: n=8, kidney: n=8, lung: n=7; simple linear regression. (B) Spike-specific CD4$^+$ T cell responder rates based on blood of non-tumor (spleen, BM) vs. tumor (liver, lung, kidney) patients. Statistically significant differences were tested with the two-sided Fisher's exact test. (C) Frequencies of overall specific blood-derived CD4$^+$ T cells, of those with a memory phenotype or expressing cytokines as depicted in patients stratified as in (B). Non-tumor: n=13, tumor: n=23. Red symbols identify vaccinated individuals with a history of SARS-CoV2 infection. Only datasets where paired blood and tissue samples were available are included, allowing comparability with the donors included in the mostly pairwise comparisons throughout the manuscript.

### Proß, Sattler, Lukassen et al. Supplemental Figure 3

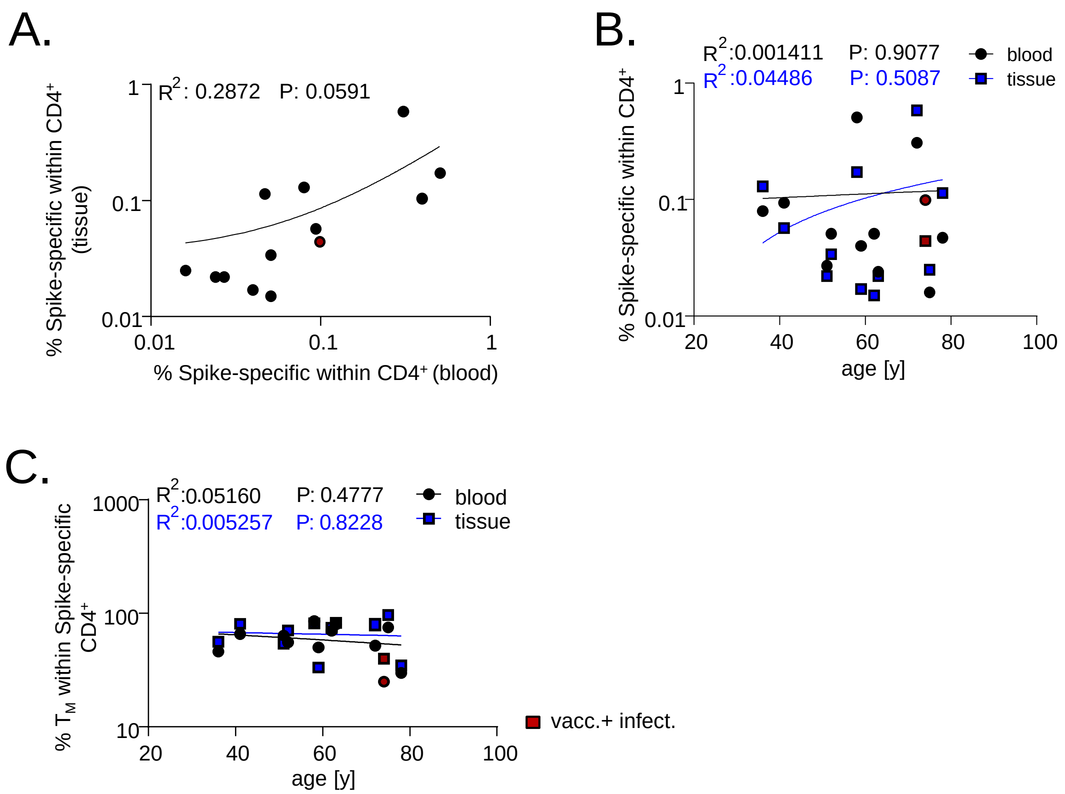


**Supplemental Figure 3. Correlation analyses for Spike-specific, lymphoid organ-derived CD4⁺ Th cells.** Correlation analyses were performed as in Figure 1H and I for frequencies of specific Th cells from lymphoid organs (BM, spleen) against those detected in blood (A) or of both against age (B). (C) Correlation analysis was performed as in Figure 2C for frequencies of antigen-specific $T_M$ from lymphoid organs. BM=10; spleen=3; simple linear regression. Red symbols identify vaccinated individuals with a history of SARS-CoV2 infection.

### Proß, Sattler, Lukassen et al. Supplemental Figure 4

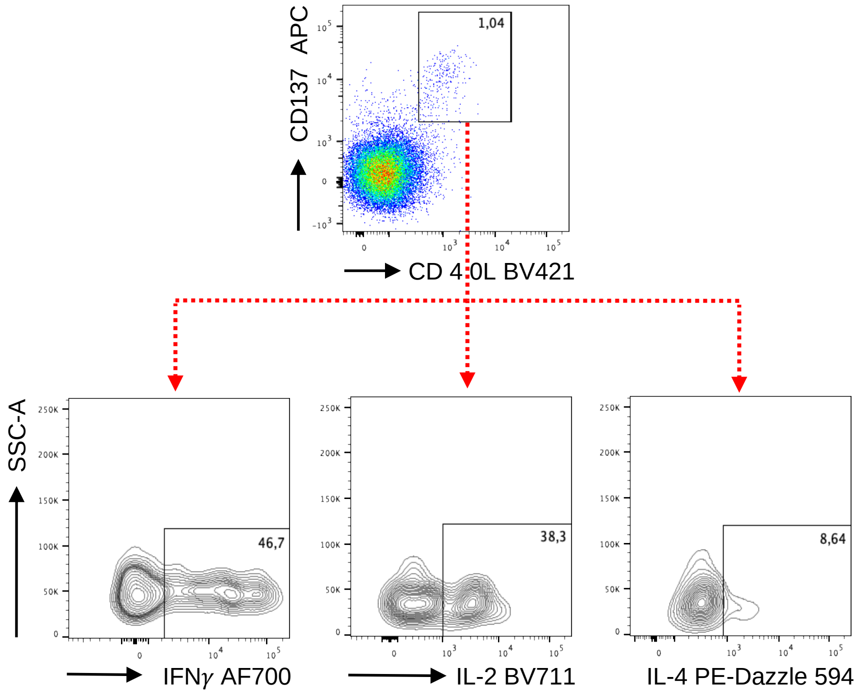


**Supplemental Figure 4. Gating strategy for identification of cytokine producing SARS-CoV-2 vaccine-specific T cells.** (A) Specific $CD4^{+}CD137^{+}CD40L^{+}$ T cells were further analyzed for expression of IFNγ, IL-2 and IL-4. Gates were applied according to the respective unstimulated controls.

### Proß, Sattler, Lukassen et al. Supplemental Figure 5

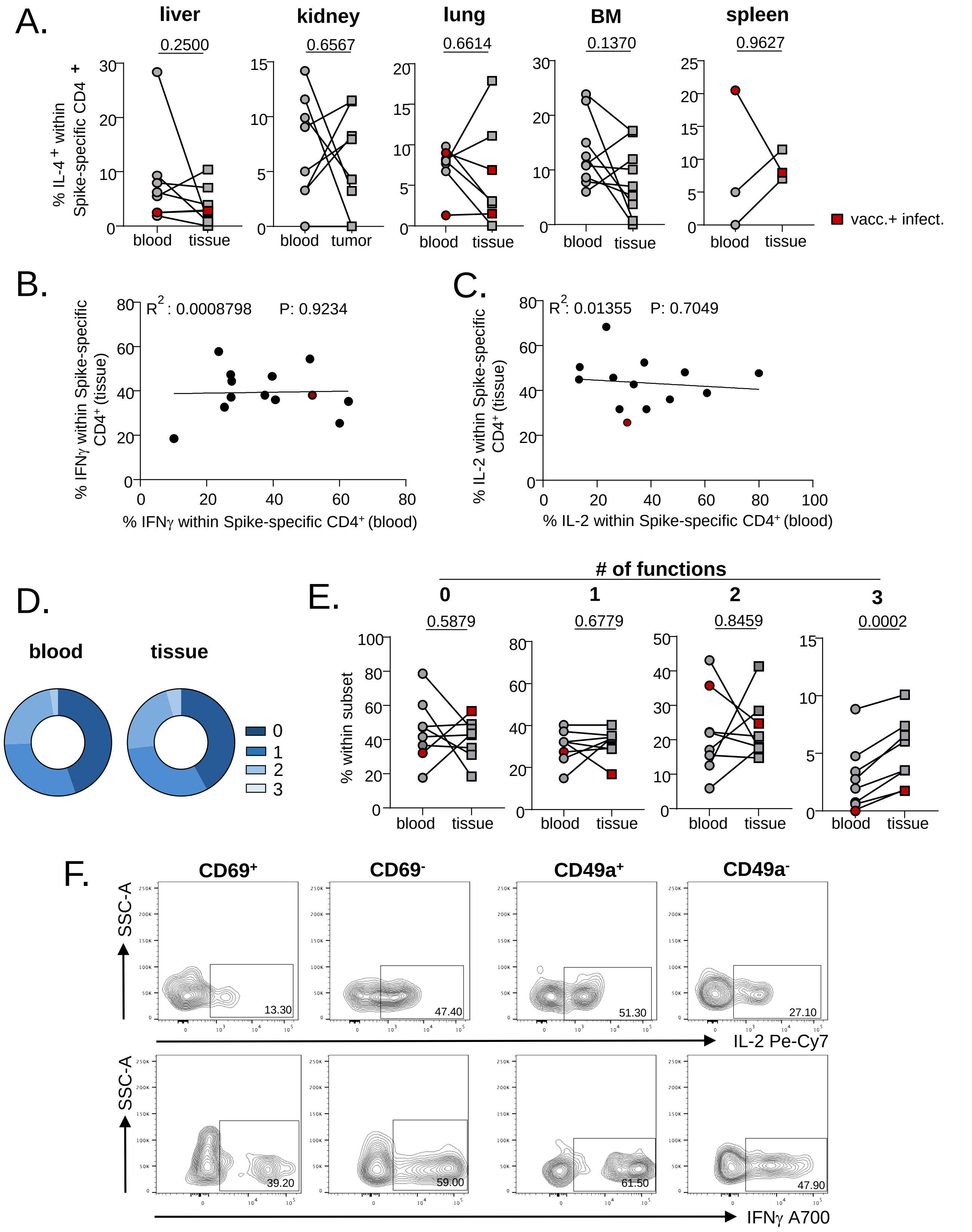


**Supplemental Figure 5. Extended functional features of Spike-specific CD4+ Th cells.** (A) FACS-based expression analysis of IL-4 in Spike-specific Th cells in paired tissue and blood samples. Liver: n=8, Wilcoxon test; kidney: n=8, paired t test; lung: n=7, paired t test; BM: n=10, Wilcoxon test; spleen: n=3, paired t test. (B) Correlation analyses were performed as in Figure 4B and C for frequencies of specific IFNγ (B) and IL-2 (C) positive Th cells from lymphoid organs (BM, spleen) against those detected in blood. BM: n=10; spleen: n=3; simple linear regression. (D and E) Polyfunctional features of specific lymphoid tissue (BM) derived Th cells as in Figures 4D and E. Statistically significant differences were tested with paired t test. (F) Exemplary plots from kidney tumor tissue illustrating differential cytokine production associated with CD69/CD49a expression, related to Figure 4F. Red symbols identify vaccinated individuals with a history of SARS-CoV2 infection.

### Proß, Sattler, Lukassen et al. Supplemental Figure 6

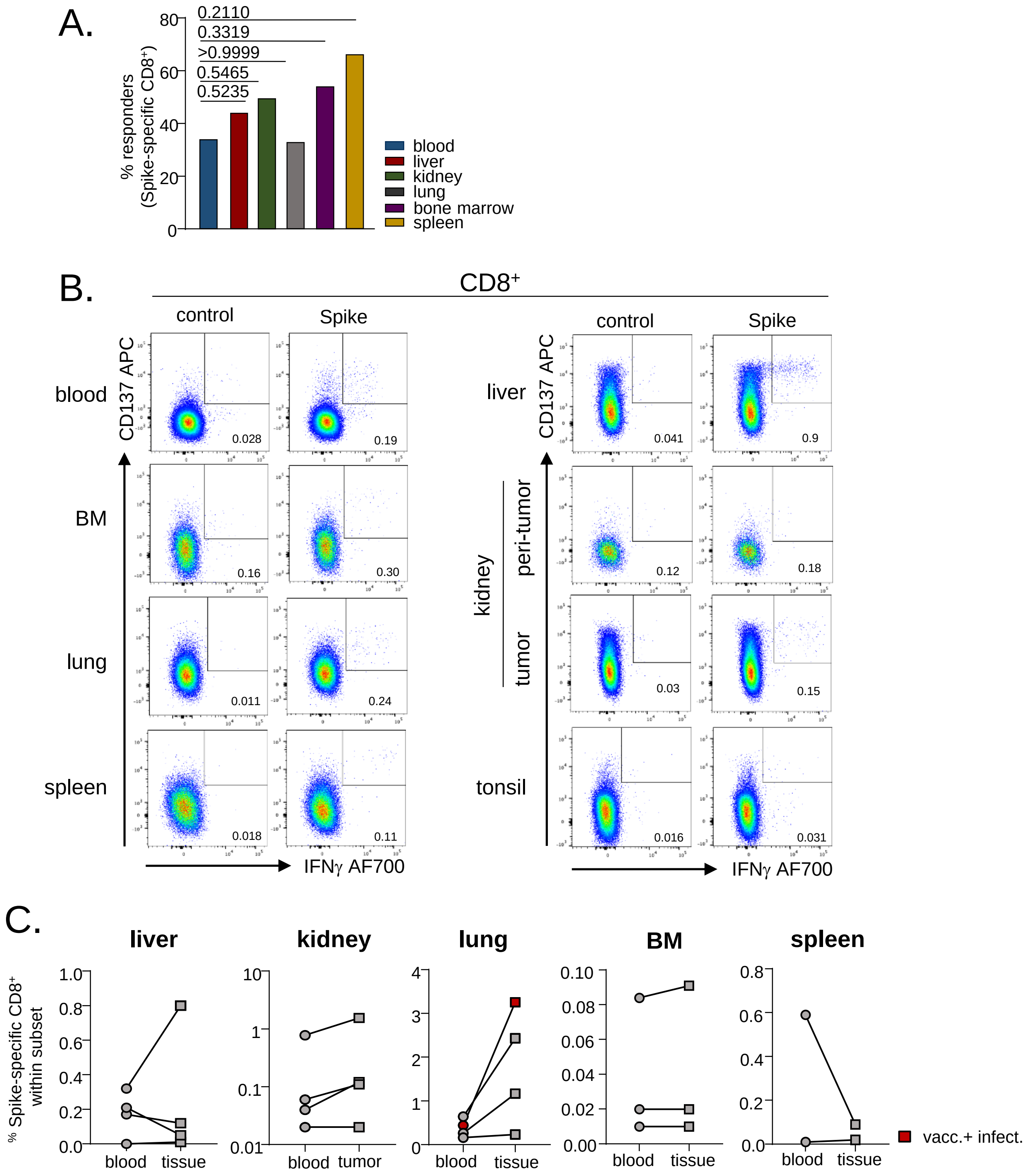


**Supplemental Figure 6. Quantification of vaccine-specific CD8⁺ T cell responses.** (A) Portions of individuals showing specific CD8⁺ T cells responses within the depicted organs. Statistically significant differences were tested with two-sided Fisher's exact test. (B) Exemplary plots and (C) summary showing vaccine-specific CD137⁺IFNγ⁺ CD8⁺ T cell quantities from the indicated organs as identified by FACS. Liver: n=4, kidney: n=4, lung: n=4, BM: n=3, spleen: n=2. Red symbols identify vaccinated individuals with a history of SARS-CoV2 infection.

### Proß, Sattler, Lukassen et al. Supplemental Figure 7

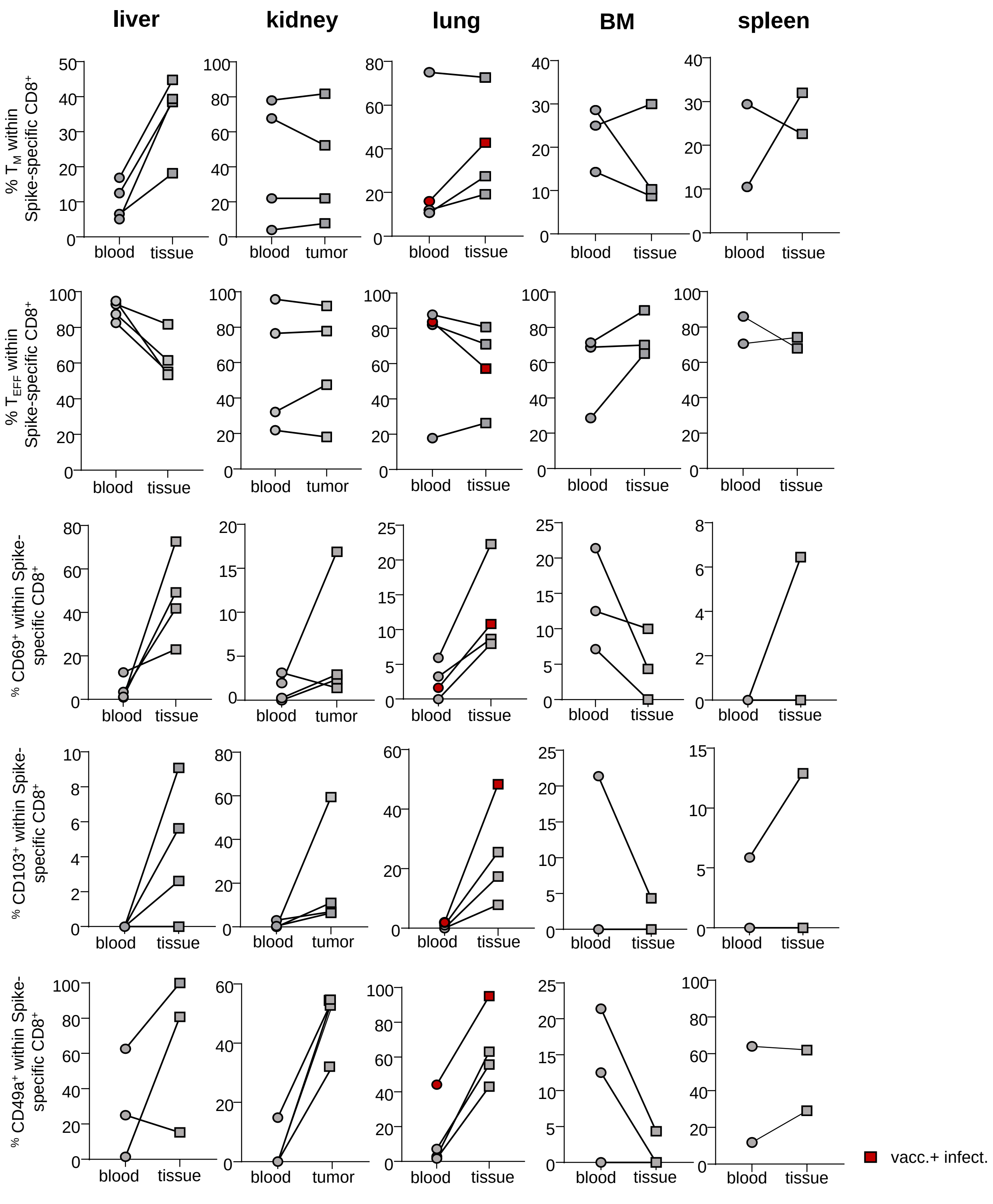


**Supplemental Figure 7. Extended phenotyping of Spike-specific CD8⁺ T cells.** Specific CD8⁺ T cells were identified as described before and further analysed for expression of memory (CD45RO, CD62L) or tissue residency/retention (CD69, CD103, CD49a) associated molecules in paired blood and organ samples as indicated. Liver: n=4, kidney: n=4, lung: n=4, BM: n=3, spleen: n=2. Red symbols identify vaccinated individuals with a history of SARS-CoV2 infection.

### Proß, Sattler, Lukassen et al. Supplemental Figure 8

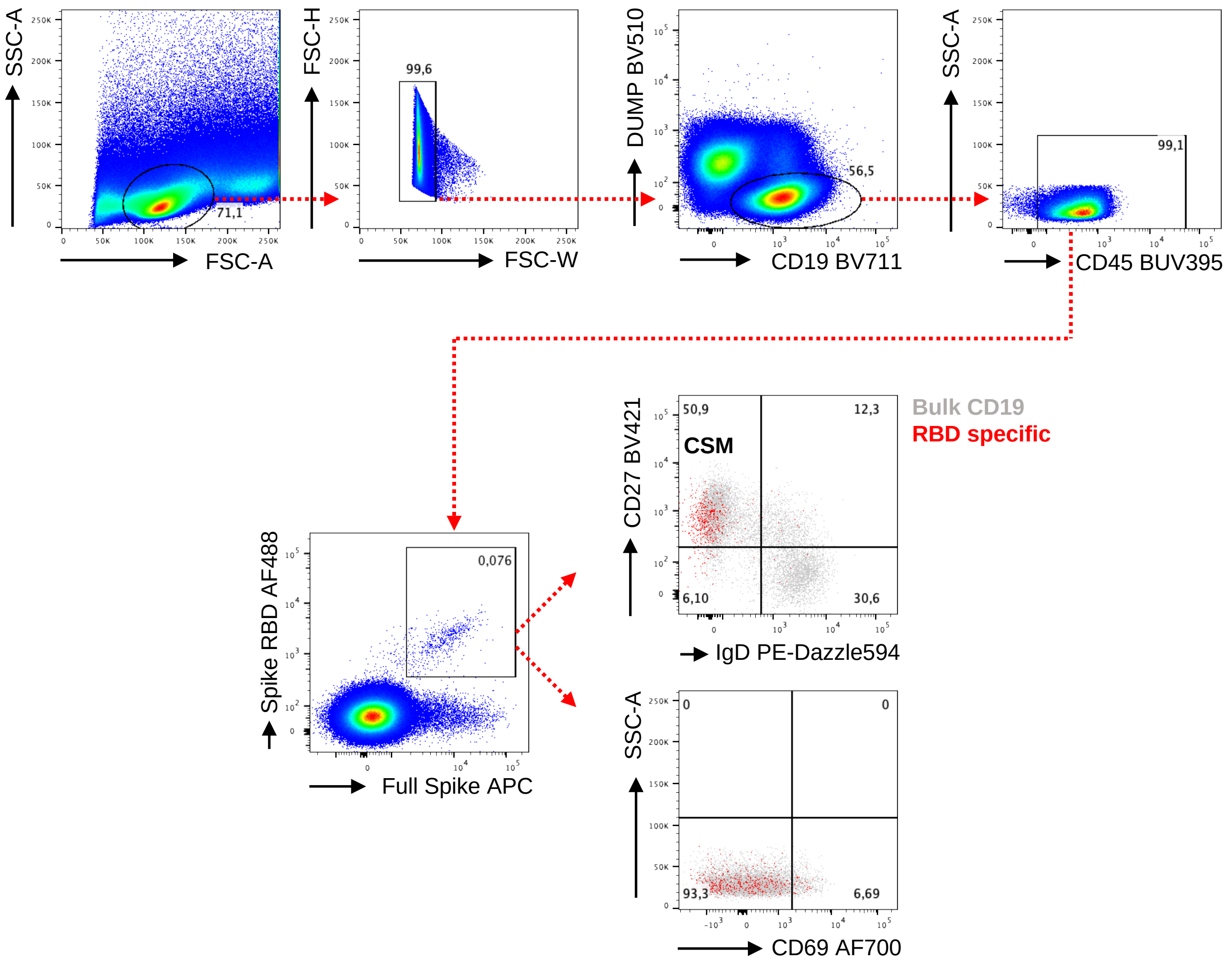


**Supplemental Figure 8. Gating strategy for assessment of SARS-CoV2-specific B cells.** Spike RBD-specific live single CD14⁻CD56⁻CD3⁻ (“dump” negative) CD19⁺ B cells were exemplarily identified in MNC from tonsil by flow cytometry based on co-staining with recombinant Spike RBD-FITC and recombinant full Spike-APC. Specific cells were further analyzed for memory differentiation (CD27, IgD), where isotype class-switched memory cells (CSM) were CD27⁺IgD⁻, or CD69 expression.

### Proß, Sattler, Lukassen et al. Supplemental Figure 9

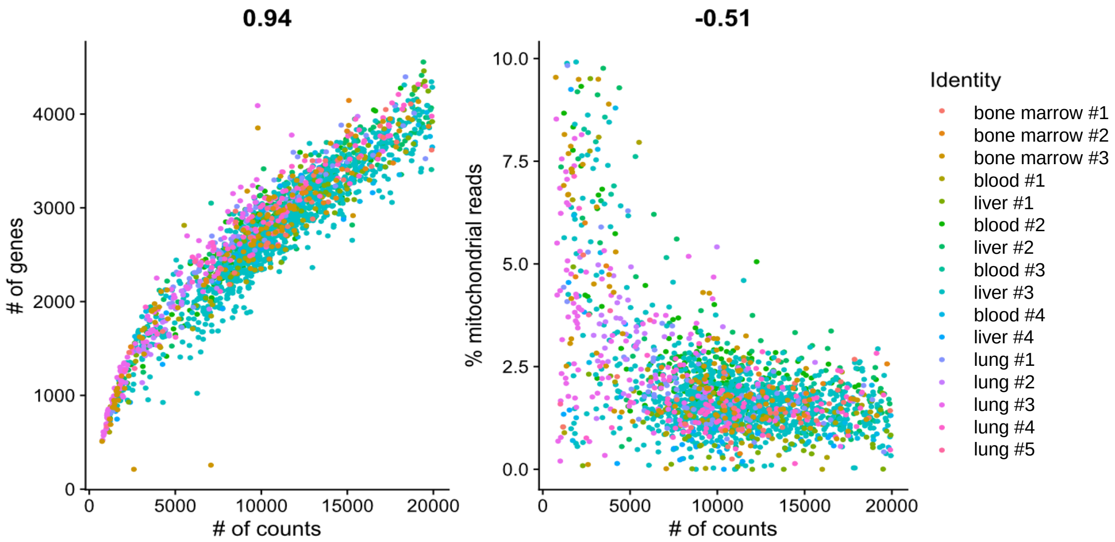


**Supplemental Figure 9.** Scatter plots of per-cell quality control metrics as depicted for single-cell RNA-Seq data. Samples are color-coded, Pearson correlation coefficients are indicated above the panels.

### Proß, Sattler, Lukassen et al. Supplemental Figure 10

**cluster 0 – „cytokine signaling“ (226 genes up-regulated)**

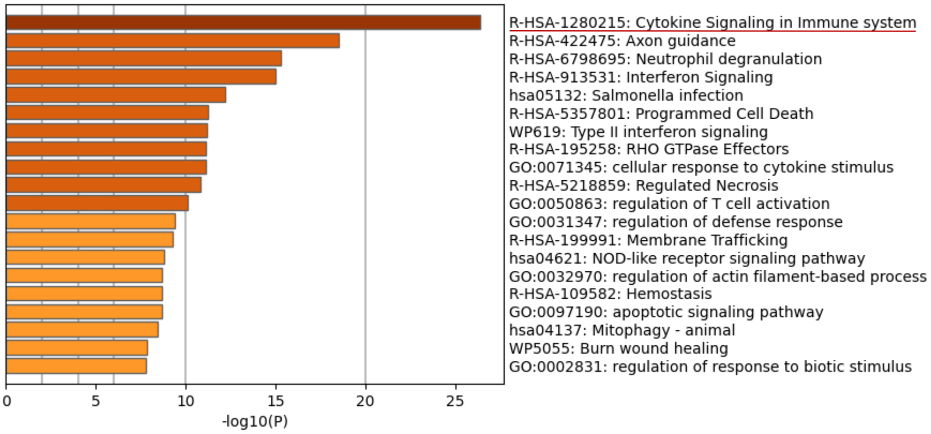


**cluster 1 - „ribosomal biogenesis“ (437 genes up-regulated)**

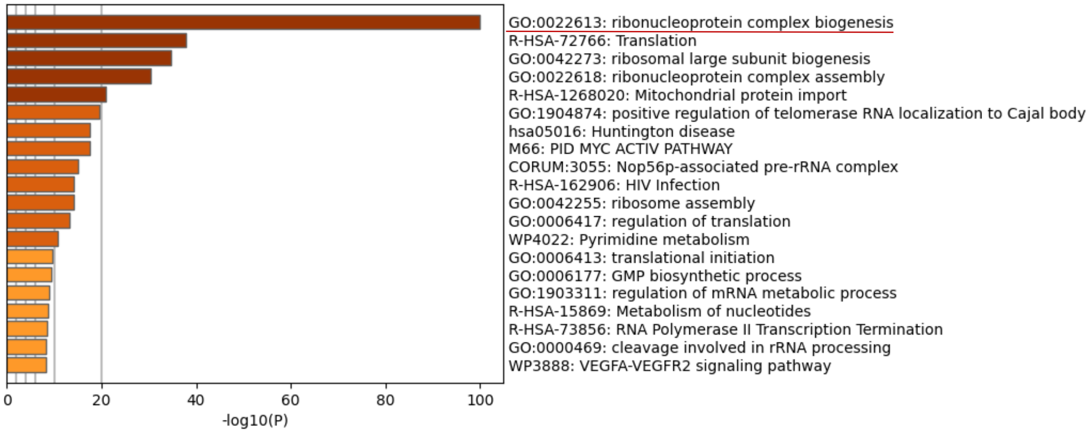


**cluster 2 – „NEAT1“ (194 genes up-regulated)**

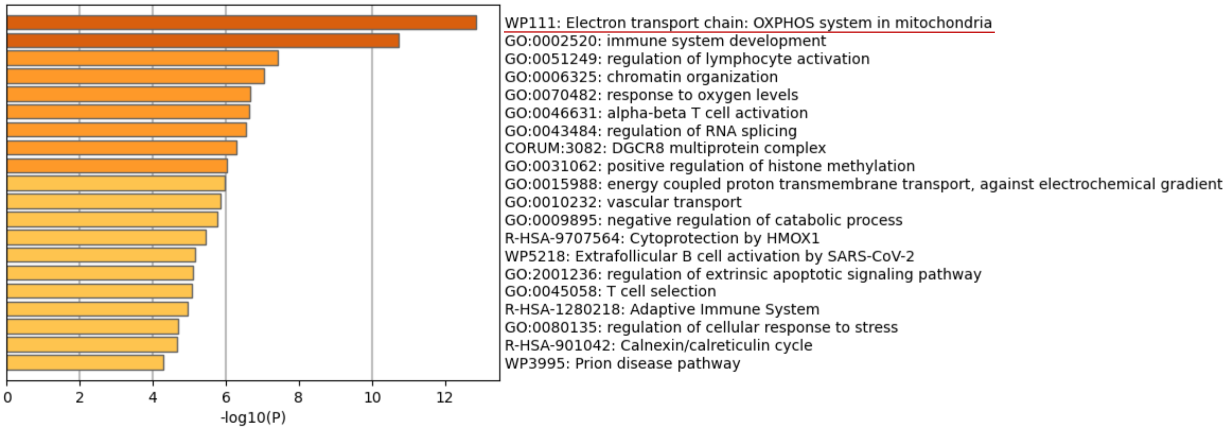


**Supplemental Figure 10**. **Pathway enrichment analysis.** Top enriched pathway analysis of transcripts enriched in cluster 0, 1 and 2. Significantly overrepresented biological processes ($p \leq 0.05$) in each cluster were plotted against Enrichment Score (calculated as -log10 p-value).

**Supplemental Table 1.** Antibodies used for T cell analysis

| Molecule | Clone | Fluorochrome | Manufacturer | Catalog Number |
|---|---|---|---|---|
| **Surface** | | | | |
| **CD3** | SK7 | PerCP/Cy5.5 | Biolegend | 344808 |
| **CD4** | SK3 | BUV395 | BD | 563550 |
| **CD8** | SK1 | APCeFluor780 | Thermo Fisher | 47-0087-42 |
| **CD14** | M5E2 | BV510 | Biolegend | 301842 |
| **CD19** | H1B19 | BV510 | Biolegend | 302242 |
| **L/D Zombie** | - | Aqua (BV510) | Biolegend | 423101 |
| **CD45RO** | UCHL1 | BV650 | Biolegend | 304232 |
| **CD49a** | TS2/7 | PE/Cy7 | Biolegend | 328311 |
| **CD62L** | DREG-56 | BV605 | BD | 304834 |
| **CD69** | FN50 | BV785 | Biolegend | 310932 |
| **CD103** | Ber-ACT8 | PE | Biolegend | 350206 |
| **Intracellular** | | | | |
| **CD137** | 4B4-1 | APC | Biolegend | 309810 |
| **CD40L** | 24-31 | BV421 | Biolegend | 310824 |
| **IFNγ** | 4S.B3 | FITC | Biolegend | 502506 |
| **IL-2** | MQ1-17H12 | BV711 | Biolegend | 500345 |
| **IL-4** | MP4-25D2 | PE-Dazzle594 | Biolegend | 500832 |

**Supplemental Table 2.** Antibodies used for B cell analysis

| Molecule | Clone | Fluorochrome | Manufacturer | Catalog Number |
|---|---|---|---|---|
| **CD3** | UCHT1 | BV510 | Biolegend | 344828 |
| **CD14** | M5E2 | BV510 | Biolegend | 301842 |
| **CD56** | 5.1H11 | BV510 | Biolegend | 362534 |
| **L/D Zombie** | - | Aqua(510) | Biolegend | 423101 |
| **CD45** | HI30 | BUV395 | BD | 563792 |
| **CD19** | SJ25C1 | BV711 | Biolegend | 363022 |
| **CD27** | M-T271 | BV421 | Biolegend | 356417 |
| **IgD** | IA6-2 | PE-Dazzle594 | Biolegend | 348240 |
| **IgG** | IS11-3B2.2.3 | PE | Miltenyi Biotec | 130-119-878 |
| **Spike RBD** | - | AF488 | R&D Systems | AFG10500-020 |
| **Full Spike** | - | Biotin | R&D Systems | BT10549-050 |
| **Streptavidin** | - | APC | Biolegend | 405207 |
| **CD69** | FN50 | AF700 | Biolegend | 310922 |

**Supplemental Table 3.** Statistics

| Figure | Statistic test |
|---|---|
| 1D | two-sided Fisher‘s exact test |
| 1E | two-sided Fisher’s exact test |
| 1F | simple linear regression analysis |
| 1G | two-tailed paired t test / Wilcoxon matched pairs signed rank test |
| 1H-J | simple linear regression analysis |
| 2B | two-tailed paired T test / Wilcoxon matched pairs signed rank test |
| 2C | simple linear regression analysis |
| 3B – 4A | two-tailed paired T test / Wilcoxon matched pairs signed rank test |
| 4B - C | simple linear regression analysis |
| 4E - F | two-tailed paired T test / Wilcoxon matched pairs signed rank test |
| 8D | Tukey‘s honest significance of differences test / ANOVA |
| Suppl. 2A | simple linear regression analysis |
| Suppl. 2B | two-sided Fisher’s exact test |
| Suppl. 2C | two-tailed unpaired T test / Mann-Whitney test |
| Suppl. 3A-C | simple linear regression analysis |
| Suppl. 5A, E | two-tailed paired T test / Wilcoxon matched pairs signed rank test |
| Suppl. 5B, C | simple linear regression analysis |
| Suppl. 6A | two-sided Fisher’s exact test |
| Suppl. 6C | two-tailed paired T test / Wlcoxon matched pairs signed rank test |
| Suppl. 9 | Pearson‘s correlation |
| Suppl. 10 | Hypergeometric test |
